# Rifaximin prophylaxis causes resistance to the last-resort antibiotic daptomycin

**DOI:** 10.1101/2023.03.01.23286614

**Authors:** A.M. Turner, L. Li, I.R. Monk, J.Y.H. Lee, D.J. Ingle, S. Portelli, N.L. Sherry, N. Isles, T. Seemann, L.K. Sharkey, C.J. Walsh, G.E. Reid, S. Nie, B.A. Eijkelkamp, N.E. Holmes, B. Collis, S. Vogrin, A. Hiergeist, D. Weber, A. Gessner, E. Holler, D.B. Ascher, S. Duchene, N.E. Scott, T.P. Stinear, J.C. Kwong, C.L. Gorrie, B.P. Howden, G.P. Carter

## Abstract

Multidrug-resistant bacterial pathogens like vancomycin-resistant *Enterococcus faecium* (VREfm) are a critical threat to human health. Daptomycin is a last-resort antibiotic for VREfm infections with a novel mode-of-action, but for which resistance has surprisingly been widely reported but unexplained. Here we show that rifaximin, an unrelated antibiotic used prophylactically to prevent hepatic encephalopathy in liver disease patients, causes cross-resistance to daptomycin in VREfm. Amino acid changes arising within the bacterial RNA polymerase in response to rifaximin exposure cause upregulation of a previously uncharacterised operon (*prdRAB*) that leads to cell membrane remodelling and cross-resistance to daptomycin through reduced binding of the antibiotic. Alarmingly, VREfm with these mutations are spread globally, making this a major mechanism of resistance. Rifaximin has been considered ‘low-risk’ for antibiotic resistance development. Our study shows this assumption is flawed and widespread rifaximin use, particularly in patients with liver cirrhosis, may be compromising the clinical use of daptomycin, a major last-resort intervention for multidrug-resistant pathogens. These findings demonstrate how unanticipated antibiotic cross-resistance can undermine global strategies designed to preserve the clinical use of critical antibiotics.

---

Antimicrobial resistance (AMR) is one of the greatest public health threats, with 1.27 million deaths being directly attributable to bacterial AMR in 2019^1^. The magnitude of this threat is therefore similar to that of malaria (558,000 global deaths in 2019)^2^, HIV (690,000 deaths in 2019)^3^, and diabetes mellitus (1.5 million deaths in 2019)^4^. Infections caused by multidrug- and extensively-drug resistant pathogens are of particular clinical concern since they are associated with frequent treatment failure and high-rates of morbidity and mortality. The preservation of last-resort antibiotics that can be used to treat these formidable pathogens is of critical importance.

*Enterococcus faecium* is one such formidable pathogen. It is a commensal of the human gastrointestinal tract that has emerged as a major cause of nosocomial infections^5^. The intrinsic antibiotic resistance of hospital-associated clones coupled with their ability to rapidly acquire additional antibiotic resistance genes makes *E. faecium* infections increasingly difficult to treat^6^. In particular, strains resistant to vancomycin, the first-line antibiotic for invasive infections, have emerged and disseminated globally due to the acquisition of transferable *van* resistance genes^7^. Consequently, *E. faecium*, which is one of the ESKAPE^8^ pathogens, has been recognised by the World Health Organization (WHO) as a ‘high priority’ bacterial pathogen^9^.

The lipopeptide daptomycin is a WHO designated last-resort antibiotic that is used ‘off-label’ to treat severe vancomycin-resistant *E. faecium* (VREfm) infections^10^. The increasing reports of daptomycin-resistant VREfm are of great clinical concern. Daptomycin resistance in clinical VREfm has been associated with the presence of specific amino acid changes in the regulatory systems LiaFSR/LiaXYZ and cardiolipin synthase Cls^11–13^. However, many daptomycin-resistant VREfm contain wild-type (WT) *liaFSR/liaXYZ* and *cls* alleles, indicating other unknown molecular pathways are involved^12,14,15^.

In Australia, high rates (15%) of daptomycin-resistant VREfm were recently reported^16^, but the data were not epidemiologically robust and the specific genetic determinants leading to resistance were not defined using molecular techniques. Accordingly, we undertook a combined genomic and phenotypic analysis to investigate the daptomycin resistance mechanisms in VREfm. Here we show that daptomycin resistance is linked with the presence of specific mutations within the *rpoB* gene, with resistance emerging *de novo* in *E. faecium* following exposure to rifaximin, a commonly prescribed antibiotic used prophylactically to prevent hepatic encephalopathy in liver disease patients^17^. The emergence of these daptomycin-associated *rpoB* mutations leads to transcriptional reprogramming in VREfm and daptomycin resistance via a mechanism that is independent of previously described systems, such as the *lia* operons or *cls.* Instead, the daptomycin-associated *rpoB* mutations cause upregulation of a previously uncharacterised genetic locus that results in cell membrane remodelling, which ultimately increases the cell surface charge and reduces daptomycin binding. Our work has uncovered a major new mechanism of daptomycin resistance in VREfm and identified rifaximin, an antibiotic considered to be low-risk for the emergence of bacterial AMR^18^, as an important driver of last-resort antibiotic resistance.

## Results

### Daptomycin resistance in Australian VREfm is polygenic and does not correlate with known resistance determinants

Daptomycin susceptibility testing was performed on VREfm isolated during two unbiased state-wide ‘snapshot’ studies undertaken for month-long periods in 2015 (n=294) and 2018 (n=423) in Victoria, Australia. The proportion of daptomycin-resistant isolates was 16.6% (n=49) in 2015 and 15.3% (n=65) in 2018. Given the high-rate of resistance observed, we expanded the study to include VREfm isolated in 2017 (n=108) and 2018 (n=173) as part of the ‘Controlling Superbugs’ study^19,20^, with 28.4% (n=80) of these isolates being resistant to daptomycin. Overall, we observed 194 (19.4%) daptomycin-resistant VREfm isolates, indicating a very high prevalence of daptomycin resistance in Victoria, Australia.

To investigate the relationship between daptomycin-resistant and daptomycin-susceptible VREfm, a maximum-likelihood phylogeny was inferred from an alignment of 6,574 core genome single nucleotide polymorphisms (SNPs) (n=1000; 998 study isolates and two controls) (Supplementary Figure 1). *In silico* multi-locus sequence typing identified 36 sequence types (STs) within the 1000 isolates; 30 of these STs included at least one of the daptomycin-resistant VREfm. Daptomycin resistance was polyphyletic, with several distinct clades. The largest clade (ST203) of daptomycin-resistant strains, accounted for 41.2% of resistant isolates (n=80 of 194) and consisted of a predominant clone during our sampling timeframe (2015 to 2018), suggestive of an expanding daptomycin-resistant lineage. The other predominant STs (ST80, ST796, ST1421, and ST1424) consisted of several groups of resistant isolates that did not cluster based on tree structure. The presence of daptomycin-resistant isolates in distinct genetic backgrounds suggested daptomycin resistance has arisen independently in VREfm on multiple occasions.

We next sought to determine the genetic determinants leading to daptomycin resistance in Australian VREfm. We systematically screened VREfm genomes for mutations in the regulatory genes *liaFSR*, *liaXYZ*, *yycFG* (*walKR*), cardiolipin synthase *(cls)*, and the division site tropomyosin locus (*divIVA*), all previously linked to daptomycin resistance^10,11,21^. Unexpectedly, there were no significant associations with daptomycin resistance and mutations in these loci (Supplementary Table 1).

### The S491F substitution in RpoB is a novel mediator of daptomycin resistance in VREfm

To identify the loci linked with daptomycin resistance, we performed a genome-wide association study (GWAS) with 1000 *E. faecium* isolates (Figure 1A). This analysis identified 142 mutations (in 73 genes) significantly (*P*<1×10^-10^) associated with daptomycin resistance (as a binary variable with a breakpoint of 8 mg L^-1^). The top five significant amino acid substitutions were (i) I274S in an uncharacterised ABC eflux protein (*P*=7.44×10^-15^), (ii) G71S in an uncharacterised permease protein (*P*=7.77×10^-14^), (iii) V288L in a mannitol dehydrogenase protein (*P*=6.08×10^-12^), (iv) S491F in RpoB, which is the RNA polymerase ß subunit (*P*=1.57×10^-13^), and (v) T634K in RpoC, which is the RNA polymerase ß’ subunit (*P*=4.40×10^-11^).

**Figure 1.**
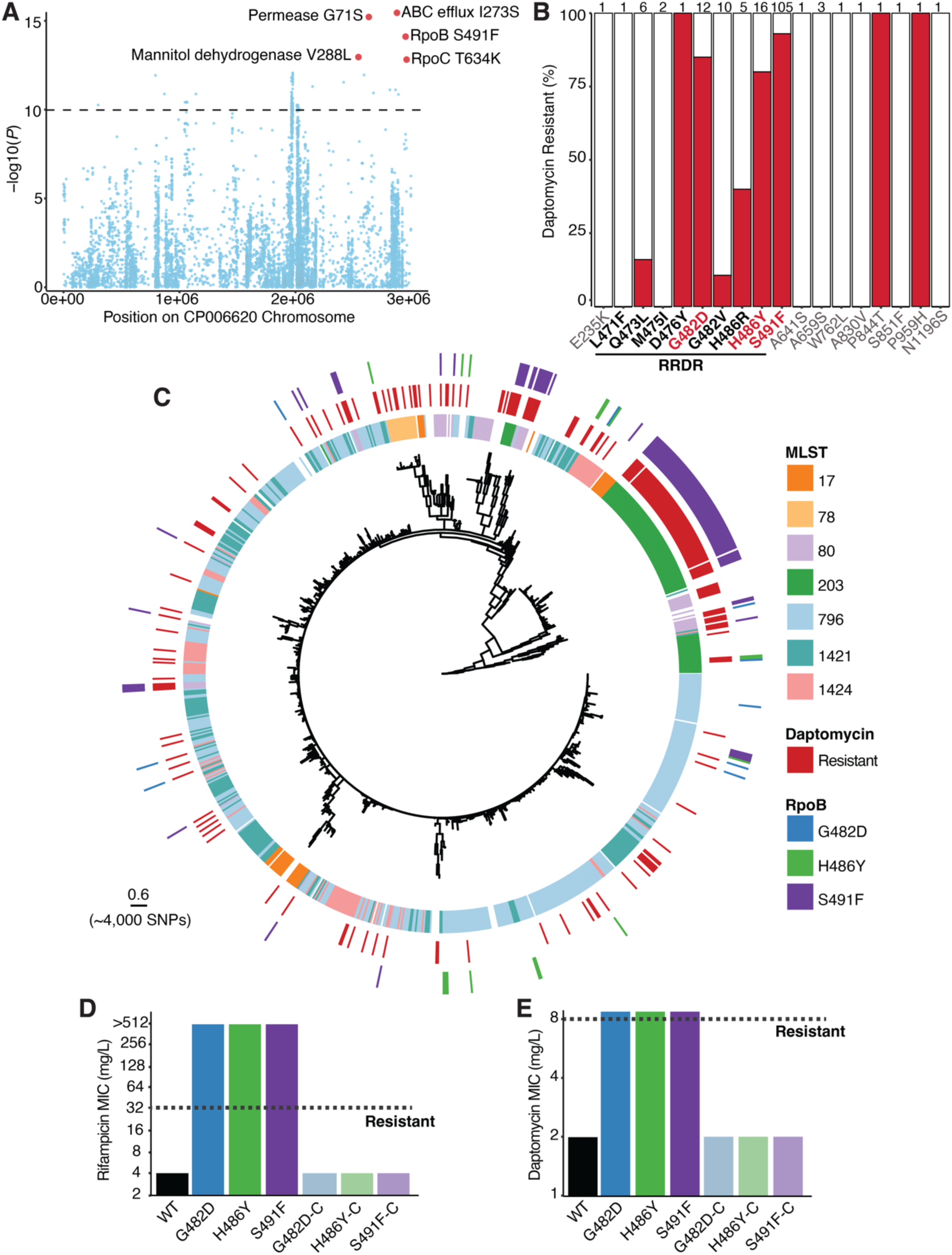
**A.** Manhattan plot of 10,530 variants, displayed by position on the reference genome and significance association with daptomycin resistance (univariate analysis using a linear mixed model). The dashed line shows the Bonferroni-corrected significance threshold. **B.** Percentage of daptomycin-resistant strains for each mutation within RpoB. The mutations within the rifampicin resistance determining region (RRDR) are shown in bold. Mutations coloured in red were associated with daptomycin resistance. The total number of strains containing each mutation is shown above each bar. **C.** Maximum-likelihood core-SNP-based phylogeny of clinical VREfm (n=1000) inferred from 6,574 SNPs, demonstrating the presence of RpoB mutations in daptomycin-resistant isolates. Overlaid are the results of *in silico* multi-locus sequence type (MLST), daptomycin phenotypic testing, and mutations associated with daptomycin resistance in RpoB. In the first circle, ST is not shown for uncommon STs (n≤5). The scale bar indicates number of nucleotide substitutions per site (top), with an approximation of SNP distance (in parentheses). **D.** Rifampicin susceptibility testing results for the WT and isogenic *rpoB* mutants and complement strains (designated with -C) (n=3). **E.** Daptomycin susceptibility testing results for the WT and isogenic *rpoB* mutants and complemented strains (designated with -C) (n=3). The MIC for each strain is shown without error bars since there was no variation between the independent biological replicates.

To test the contribution of these amino acid substitutions on daptomycin resistance we engineered each mutation into a clinical daptomycin-susceptible strain of VREfm (ST796). Introduction of the I274S substitution in the ABC eflux protein, G71S substitution in the permease protein, V288L substitution in the mannitol dehydrogenase protein, or T634K substitution within RpoC had no impact on daptomycin susceptibility. However, introduction of the S491F substitution within RpoB resulted in a daptomycin-resistant phenotype (4-fold increase in daptomycin MIC, from 2 mg L^-1^ to 8 mg L^-1^). Interestingly, every clinical strain harbouring the RpoB S491F substitution (n=105, spanning different *E. faecium* genotypes) also contained the RpoC T634K substitution (n=105), with no strains containing the RpoC T634K in isolation. Introduction of the RpoC T634K substitution into the RpoB S491F mutant background did not affect daptomycin susceptibility (MIC still 8 mg L^-1^), suggesting this mutation does not play a direct role in the daptomycin resistance phenotype and instead may be compensatory for negative fitness effects associated with the RpoB S491F substitution. In a competition assay (WT versus RpoB S491F or WT versus RpoB S491F/RpoC T634K), the RpoB S491F substitution posed a substantial fitness cost, with a significant (*P*<0.0001) shift to the WT population (Supplementary Figure 2). However, in the double RpoB S491F/RpoC T634K mutant the population dynamics remained stable, with no significant differences compared to inoculation observed (Supplementary Figure 2). These data suggest the RpoC T634K substitution is compensatory for the negative fitness effects of RpoB S491F, a hypothesis consistent with studies in *Mycobacterium tuberculosis*^22^ and *Escherichia coli*^23^.

### Different substitutions within the rifampicin resistance determining region of RpoB result in daptomycin resistance in VREfm

We next assessed if other RpoB substitutions were associated with daptomycin resistance (Figure 1B). We observed resistance in 81.3% of VREfm strains (n=16) with a G482D substitution and 83.3% of VREfm strains (n=12) with a H486Y substitution. Isolates harbouring these mutations were spread across the phylogenetic tree, indicative of multiple independent acquisitions (Figure 1C). No putative compensatory mutations in the RNA polymerase genes were identified. Clonal expansion was also observed for a dominant, daptomycin-resistant clone (ST203) containing the S491F substitution. Daptomycin-resistant isolates from this ST203 lineage were identified across 10 geographically distinct hospital networks, indicating they were not part of a singular hospital outbreak (Supplementary Figure 3).

The G482D, H486Y, and S491F substitutions were located within the predicted rifampicin-resistance determining region (RRDR) of RpoB (spanning amino acids 467 to 493). Rifampicin susceptibility testing (rifampicin being a marker of rifamycin resistance^24^) confirmed all isolates had high-level rifampicin resistance (n=169, median MIC 256 mg L^-1^), while control isolates containing the WT RRDR displayed a median MIC of 8 mg L^-1^ (n=169, Supplementary Figure 4).

To test if the G482D and H486Y substitutions in RpoB also led to rifamycin and daptomycin resistance we engineered isogenic mutants with these substitutions in our rifamycin-susceptible, daptomycin-susceptible, clinical strain of VREfm (ST796). Introduction of the G482D, H486Y, or S491F RpoB substitutions resulted in a 7-fold decrease in rifampicin susceptibility compared to the WT strain, leading to high-level rifampicin resistance (>512 mg L^-1^) (Figure 1D). The introduction of the G482D or H486Y RpoB substitutions also resulted in a daptomycin-resistant phenotype (4-fold increase in daptomycin MIC, from 2 mg L^-1^ to 8 mg L^-1^) (Figure 1E). Reverse allelic exchange with the WT *rpoB* allele resulted in susceptibility to rifampicin (MIC of 4 mg L^-1^) and daptomycin (MIC of 2 mg L^-1^), confirming that G482D, H486Y, and S491F caused rifamycin resistance and daptomycin cross-resistance.

### The G482D, H486Y, and S491F RpoB substitutions are common in international VREfm strains

To determine if the RpoB substitutions associated with daptomycin resistance observed in Australian VREfm were representative of other *E. faecium* globally, we analysed publicly available VREfm sequence data (n=3,476 international and n=1,000 Australian) (Figure 2A). The majority (n=3,378) of these VREfm were healthcare-associated, with 98 isolates from animal origin. Of all the isolates analysed, 630 (14.1%) carried an amino acid substitution in the RRDR of RpoB, with the S491F substitution being the most common, present in 77.9% (n=461) of isolates with a RRDR RpoB substitution (Figure 2B). The S491F substitution was identified in isolates from 20 countries and across 21 different STs (Figure 2B and Supplementary Figure 5A). Importantly, five VREfm (from three genetic backgrounds and four countries) harbouring the S491F substitution also contained *cfr(B)* (n=4) or *poxtA* (n=1) linezolid resistance genes, suggesting near pan-resistant strains of VREfm have emerged. The G482D and H486Y substitutions were also commonly identified, found in 6.8% (n=43) and 11.6% (n=73) of strains with the RpoB substitution, respectively. The G482D and H486Y substitutions were also observed globally (7 and 10 countries, respectively) and across different STs (9 and 22, respectively) (Figure 2B and Supplementary Figure 5A). These data indicated the RpoB substitutions conferring resistance to rifamycins and cross-resistance to daptomycin are globally prevalent. We also observed a significant association between RRDR RpoB substitutions and healthcare-associated VREfm (*P*<0.001; Fisher’s exact test) (Supplementary Figure 5B)^25^, suggesting the identified RpoB substitutions are enriched within the healthcare setting.

**Figure 2.**
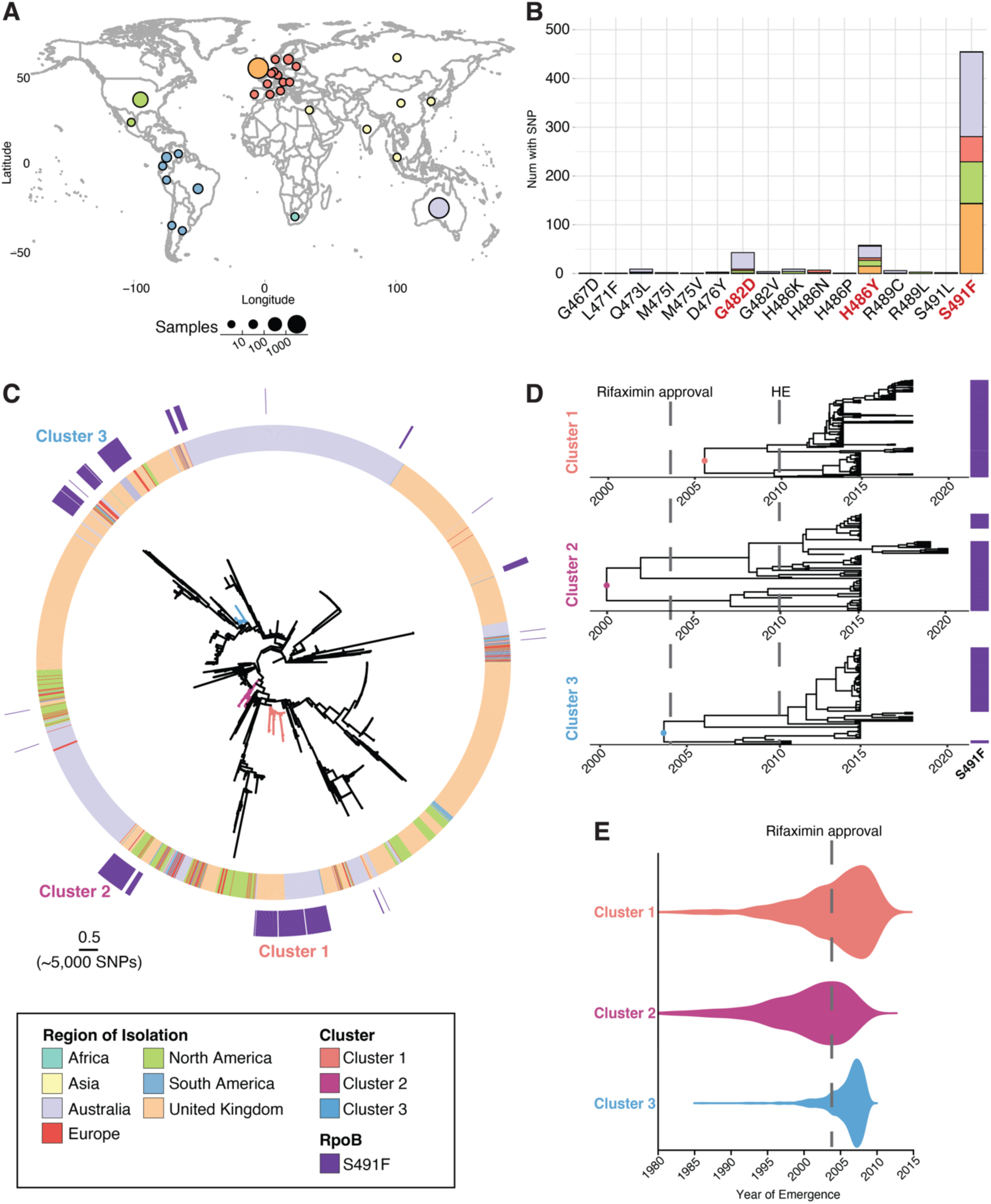
**A.** Map of 4,476 VREfm genomes included in this study. Circle size corresponds to total number of genomes (binned into categories), and colour corresponds to region of isolation. Country coordinates used are the country centroid position. **B.** The frequency of various RpoB mutations within the rifampicin resistance determining region (RRDR) in 4,476 VREfm genomes, sampled from 43 MLSTs. Bars are coloured by the number of isolates from each region of isolation containing the mutation. The identified daptomycin resistance associated mutations are coloured in red. **C.** Maximum-likelihood, core-SNP-based phylogeny for 4,476 VREfm inferred from 9,277 core-genome SNPs, demonstrating the presence of the S491F RpoB mutation in international VREfm. Overlaid is the region of isolation for each strain and the presence of the S491F mutation. The coloured branches indicate the three VREfm clusters identified with cgMLST used as input for Bayesian evolutionary analyses. The scale bar indicates number of nucleotide substitutions per site (top), with an approximation of SNP distance (in parentheses). **D.** Bayesian phylodynamic analyses showing the maximum-clade credibility (MCC) trees of the three VREfm clusters with the timing of emergence for each lineage. The median substitution rate was similar for Cluster 1 and Cluster 2, at 9.7×10-7 [95% highest posterior density (HPD) 6.88×10-7 – 1.24×10-6] and 1.25×10-6 (95% HPD 7.68×10-7 – 1.74×10-6) respectively, but slightly faster for Cluster 3 at 3.86×10-6 (95% HPD 2.23×10-6 – 5.69×10-6). Cluster 1 (n=219) was inferred from a core alignment of 1,869,554 bp containing 329 SNP sites; Cluster 2 (n=85) was inferred from an alignment of 1,524,024 bp containing 541 SNP sites; and Cluster 3 (n=68) was inferred from an alignment of 1,860,780 bp containing 764 SNP sites. The time in years is given on the *x* axis. The presence of the S491F RpoB trait for each isolate is shown in purple. Overlaid onto the MCC trees is the first instance of FDA approval for rifaximin (2004) and for hepatic encephalopathy (HE) (2010). **E.** Violin plots for the most recent common ancestor (MCRA) for each cluster, representing when the RpoB S491F mutation first emerged, with 95% HPD intervals – 2006 (HPD 1993 – 2012) for Cluster 1, 2000 (HPD 1989 – 2008) for Cluster 2, and 2004 (HPD 2001 – 2010) for Cluster 3. Overlaid onto violin plots is the FDA approval date for rifaximin (2004).

### Phylodynamics show the S491F RpoB substitution emerged within the VREfm population in the same period as the clinical approval of rifaximin

We used phylodynamic analyses to estimate the emergence date of VREfm harbouring the S491F substitution globally. Within the genomes of Australian VREfm isolates, we observed the expansion of a dominant ST203 clone from 2015 to 2018 that carried the S491F substitution (Figure 1C). Since this clone comprised VREfm carrying the *vanA* resistance operon, we genome sequenced all historical *vanA*-VREfm from our public health laboratory (n=229), which consisted of every *vanA*-VREfm isolate collected from 2003 to 2014, to increase the potential to detect a molecular clock signal. Using comparative genomics, we contextualised all Australian isolates (n=1,229) with the international data (n=3,476) and identified three distinct clusters containing the RpoB S491F substitution (Figure 2C). The same ST203 clone formed the largest cluster (Cluster 1; n=219 taxa), consisting of isolates from Australia and United Kingdom. Cluster 2 (n=85 taxa) consisted of ST80 and ST78 isolates from Australia, Europe, South America, the United Kingdom, and the United States while Cluster 3 (n=68 taxa) consisted of ST80 isolates from Australia, Europe, and the United Kingdom.

To model the evolutionary trajectories of these clusters, we used core-genome SNP diversity and year of isolation (Figure 2D and Supplementary Figure 6). The substitution rate (the number of expected nucleotide substitutions per site per year) was consistent with other estimates for healthcare-associated VREfm^25–28^ (Figure 2D). The year of emergence for the most recent common ancestor (MRCA) of each cluster with S491F was around 2006 (Figure 2D), a time period that coincides with the first clinical use of rifaximin. Since rifampicin was approved for clinical use in the United States in 1971^29^, several decades before the estimated emergence of the S491F-containing VREfm strains, we considered this rifamycin unlikely to have played a major role in the spread of resistance. Analysis of the three maximum clade credibility (MCC) trees (Figure 2D) indicated each *E. faecium* lineage has continued to expand since its emergence, consistent with the growing use of rifaximin globally, in particular since 2010 when it was approved for the prevention of hepatic encephalopathy^17^ (Figure 2D-E). It is also noteworthy that in all three VREfm clusters, the S491F substitution has been stably maintained after its acquisition, suggestive of on-going selective pressure acting on the bacterial population, as *rpoB* mutations usually carry a fitness cost (Supplementary Figure 7A-C)^22^. These data show the S491F substitution emerged within the VREfm population on at least three occasions since the early 2000s, with the predicted dates of emergence closely correlated with the clinical introduction of rifaximin.

### Patients receiving rifaximin are more likely to carry VREfm with daptomycin resistance RpoB substitutions than patients that have not received rifaximin

Rifaximin is a non-absorbable oral agent with direct antimicrobial activity in the gastrointestinal tract, predominately prescribed to prevent recurrent hepatic encephalopathy in patients with liver cirrhosis^17,30^. This patient cohort is high-risk for VREfm gastrointestinal colonisation^31^. Since the Bayesian phylodynamic analyses highlighted a correlation between the S491F RpoB substitution in VREfm and use of rifaximin, we hypothesised rifaximin use in VREfm colonised patients may be associated with the presence of substitutions in RpoB and, therefore, daptomycin resistance. To test this hypothesis, we assessed the association between rifaximin exposure and daptomycin-resistant VREfm in a retrospective patient cohort at a quaternary referral healthcare centre in Melbourne, Australia. The *E. faecium* isolates from patients with current or prior rifaximin exposure – “rifaximin” group – and without prior exposure (“control” group) were assessed a) genomically for the RpoB amino acid substitutions (G482D, H486Y, and S491F) that result in daptomycin resistance and b) phenotypically for daptomycin resistance, by study investigators blinded to patient details and the exposure groups. The VREfm isolates were phylogenetically distributed and both rifaximin and control group isolates were representative of the *E. faecium* strains identified in the state-wide snapshot survey (Figure 3A)^32^. Isolates that were genomically clustered and likely to represent patient-to-patient transmission were excluded from the analysis (see Methods). Genetically distinct VREfm from 212 individual patients collected between 2014-2022 were included in the analysis: 96 in the “rifaximin” exposed group and 116 in the unexposed “control” group. Overall patient demographics of the cohort were balanced between the two groups with the exceptions of age (*P*<0.001) and overall Charlson Comorbidity Index score (*P*<0.001), which was driven by a significantly greater proportion of patients with chronic liver disease in the rifaximin group (*P*<0.001) (Supplementary Table 2). As expected, there was also significantly greater recent exposure to rifaximin (within the last 90 days prior to VREfm sample collection) in the rifaximin group (*P*<0.001). Notably, only a single patient in the study had received recent daptomycin.

**Figure 3.**
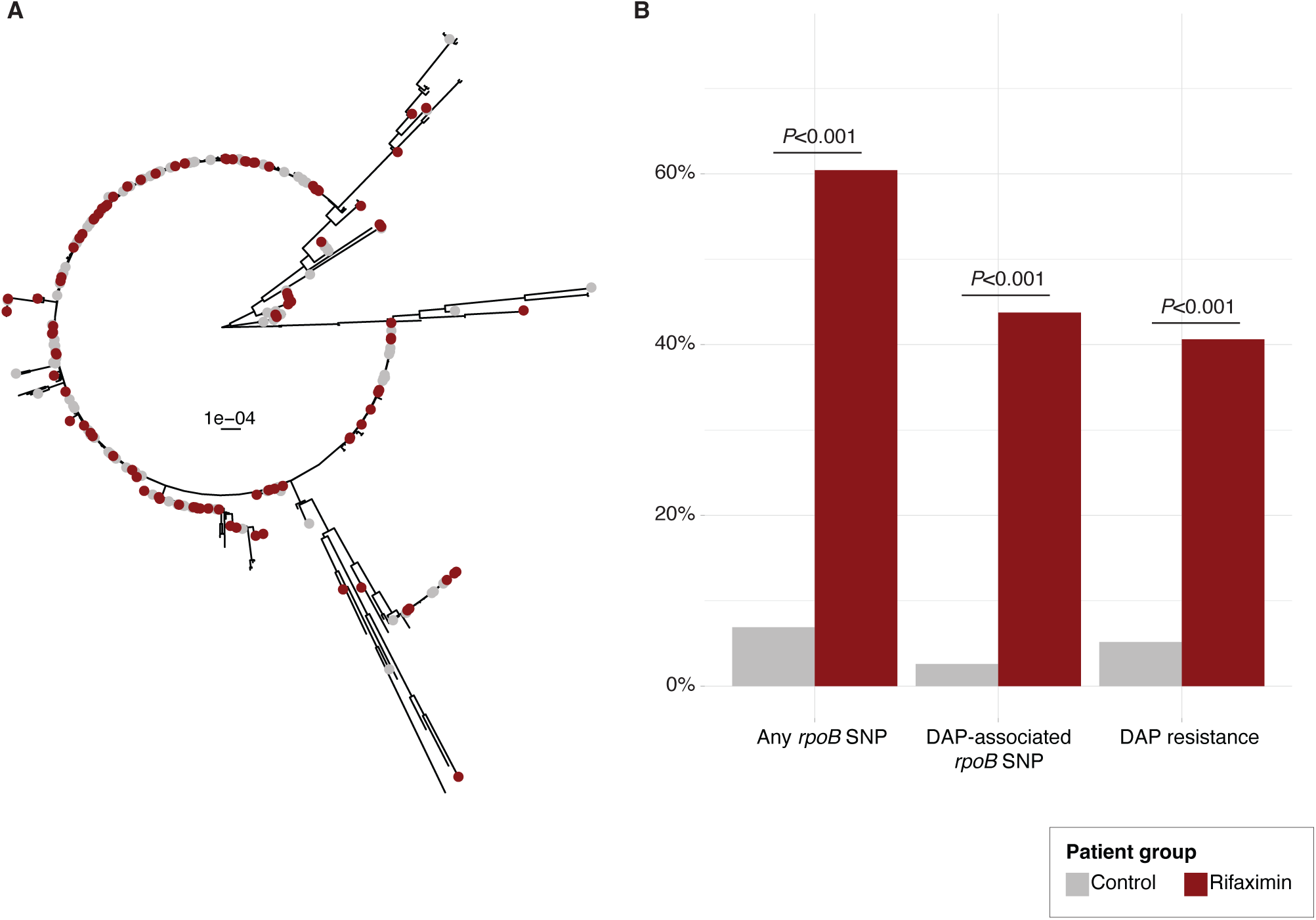
**A.** Maximum-likelihood, core-SNP-based phylogeny for VREfm inferred from 14,420 core-genome SNPs, demonstrating which isolates were from the “control” or “rifaximin” groups. The scale bar indicates number of nucleotide substitutions per site. **B.** Summary of the percentage of VREfm with any *rpoB* SNP, daptomycin (DAP)-associated *rpoB* SNP (S491F, G482D and H486Y), or DAP resistance in patients in the “control” or “rifaximin” groups. Data was analysed using a Fisher’s exact test.

Among the VREfm obtained from the entire (n=212) cohort, recent exposure to rifaximin was significantly associated with the presence of RpoB substitutions (*P*<0.001), including substitutions associated with daptomycin resistance (either G482D, H486Y, or S491F; *P*<0.001), and was significantly associated with carriage of daptomycin-resistant VREfm (P*<*0.001) (Figure 3B, Supplementary Table 3). More importantly, after adjusting for potential confounders, recent rifaximin exposure remained an independent predictor of these daptomycin resistance RpoB substitutions (OR 8.69; 95% CI 2.95-30.84; *P*<0.001) and daptomycin-resistant VREfm (OR 6.47; 95% CI 2.34-20.80; *P*<0.001) among the overall cohort of patients (Supplementary Table 4A-B). Given almost all patients who received rifaximin had underlying chronic liver disease (with hepatic encephalopathy being the predominant indication for rifaximin prescribing in Australia), we assessed the association between rifaximin and daptomycin resistance first within the subgroup of patients with liver disease (Supplementary Tables 5A-C), and then second, in an independent cohort of patients without liver disease from the University Medical Center, Regensburg, Germany. These patients were undergoing haematopoietic stem cell transplantation (HSCT), where rifaximin is used for antimicrobial prophylaxis (Supplementary Tables 6A-C). The significant association between rifaximin exposure and daptomycin resistance RpoB substitutions was again identified in both patient groups – a) patients with chronic liver disease (*P*=0.001) and b) HSCT patients without liver disease (*P*<0.001) (Supplementary Figure 10 and Supplementary Tables 5C and 6C). Among the patients with liver disease colonised with VREfm, recent rifaximin was an independent predictor of daptomycin-resistant VREfm (OR 4.37; 95% CI 1.70-12.84; *P*=0.004). We conducted several other sensitivity analyses to verify the robustness of these associations and assess for other confounders, including a subgroup analysis that excluded patients who received rifaximin from the analysis (Supplementary Table 7), but these did not identify any other associations between daptomycin resistance and underlying disease or antibiotic exposure.

There were six patients from the “control” group with daptomycin-resistant VREfm – three (50%) of these had isolates with no mutations in *rpoB*, suggesting an alternate mechanism for resistance, while the remaining three patients had previously been admitted to the liver ward for inpatient care, suggesting they may have acquired daptomycin-resistant VREfm through healthcare-associated transmission.

While only one representative VREfm isolate from each patient was included in the above analysis, the *de novo* emergence of daptomycin-resistant VREfm carrying the G482D RpoB substitution was observed in one patient from whom multiple isolates were collected during rifaximin therapy, consistent with rifaximin exposure driving the *de novo* emergence of daptomycin-resistant VREfm.

Therefore, in agreement with our laboratory observations, these clinical data confirm a strong clinical association between recent rifaximin exposure and *rpoB* substitutions linked to daptomycin resistance among patients colonised with VREfm, suggesting exposure to rifaximin could drive daptomycin-resistant VREfm.

### Rifaximin exposure leads to daptomycin-resistant VREfm in a murine model of gastrointestinal colonisation

We next used a mouse VREfm gastrointestinal colonisation model to understand if rifaximin exposure caused *de novo* emergence of *rpoB* mutations that confer cross-resistance to daptomycin. Mice were colonised with a daptomycin-susceptible (MIC 2 mg L^-1^) clinical VREfm isolate (Aus0233) containing a WT *rpoB* allele before being administered a human-equivalent dose of either rifaximin, rifampicin, daptomycin, or vehicle (Figure 4A and Supplementary Figure 9). Rifampicin was chosen as a comparison since it is a rifamycin used in clinical practice. After 7 days of rifamycin treatment, we observed rifamycin-resistant VREfm in significantly more mice receiving rifampicin (80% of mice) or rifaximin (90% of mice) than in mice that received daptomycin (0% of mice) (*P*<0.0001 and *P*<0.0001; unpaired *t*-test) or vehicle (0% of mice) (*P*<0.0001 and *P*<0.0001; unpaired *t*-test) (Figure 4B).

**Figure 4.**
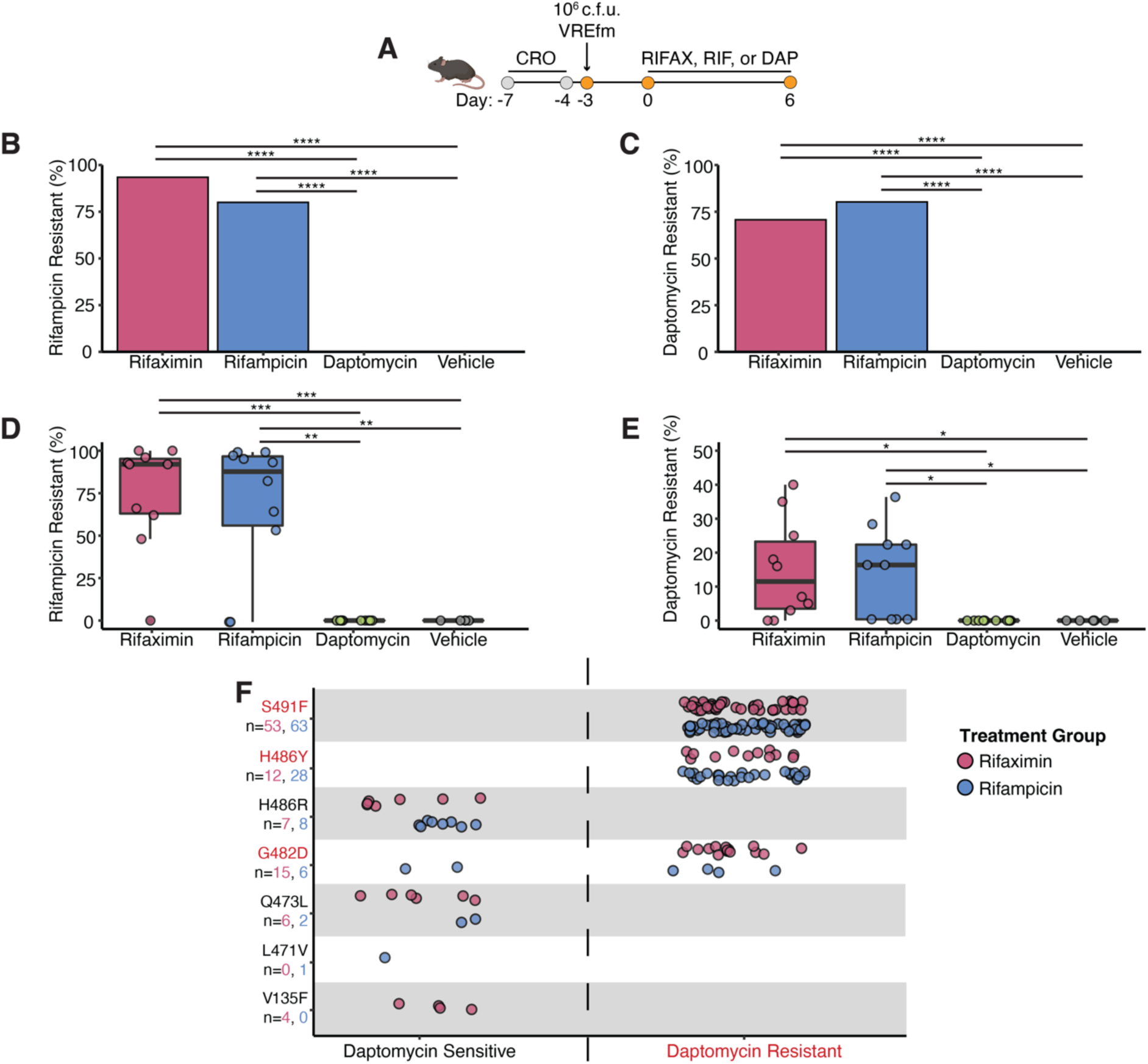
**A.** Timeline of the mouse experiment. VREfm-colonised mice (n=5 for vehicle, n=10 for rifampicin, n=10 for rifaximin, n=10 for daptomycin) received a human-equivalent dose of vehicle, rifampicin, or rifaximin (twice per day for rifaximin) for 7 days by oral gavage or subcutaneous injection with daptomycin for 7 days. CRO=ceftriaxone; DAP=daptomycin; RIFAX=rifaximin; RIF=rifampicin. Figure to scale. **B.** Percentage of total mice with rifampicin-resistant VREfm strains **C.** or daptomycin-resistant VREfm strains after 7 days of antibiotic treatment. **D.** Percentage of VREfm from each mouse (n=50 colonies per mouse) that were resistant to rifampicin **E.** or daptomycin after 7 days of antibiotic treatment. Points represent an individual mouse. Percentage was calculated from rifampicin or daptomycin MIC values (either resistant or susceptible) from 50 VREfm colonies isolated from each mouse. Boxes represent the median and interquartile range for each group. **F.** Overview of the RpoB mutations identified in the rifampicin-resistant colonies. Each point represents a single colony. Isolates are separated by each RpoB mutation and grouped into either daptomycin-susceptible or daptomycin-resistant. The RpoB mutations coloured in red had an association with daptomycin resistance; n values represent the number of isolates containing each mutation for rifaximin and rifampicin, respectively. \**P* < 0.05, \*\**P* < 0.01, \*\*\**P* < 0.001, \*\*\*\**P*<0.0001; unpaired *t*-test (vehicle versus rifampicin or vehicle versus rifaximin and rifampicin verses daptomycin or rifaximin verses daptomycin).

For each mouse, we then determined the percentage of individual VREfm isolates that were rifamycin-resistant or daptomycin-resistant. There were significantly more rifamycin-resistant VREfm isolated from mice receiving rifaximin or rifampicin than mice receiving the vehicle control (*P*<0.0001 and *P*<0.0001; unpaired *t*-test) or daptomycin (*P*<0.001 and *P*<0.001; unpaired *t*-test) (Figure 4D). Similarly, there was significantly more daptomycin-resistant VREfm in mice receiving rifaximin or rifampicin than vehicle control (*P*<0.05 and *P*<0.05; unpaired *t*-test) or daptomycin (*P*<0.05 and *P*<0.05; unpaired *t*-test) (Figure 4E). We estimated that daptomycin-resistant VREfm accounted for between 0-41% of the gastrointestinal VREfm population in mice given rifaximin and 0-36% in mice given rifampicin, demonstrating conclusively that rifamycin administration drives the emergence of VREfm with resistance to rifamycins and cross-resistance to daptomycin. Notably, no daptomycin-resistant VREfm were isolated from mice receiving daptomycin, in agreement with prior research^35^.

We then performed WGS on 150 randomly selected VREfm isolates from each group (rifaximin or rifampicin, n=300 total) to identify all mutations present in the rifamycin-resistant isolates. This collection consisted of 100 rifaximin or rifampicin-resistant isolates taken following the last day of treatment and 50 isolates from each group collected prior to rifamycin administration. No substitutions in RpoB were identified in any *E. faecium* isolate taken prior to rifaximin or rifampicin exposure. However, after exposure to either rifaximin or rifampicin, VREfm carrying mutations within *rpoB* were commonly identified. The S491F substitution was most prevalent (n=53 [rifaximin] and n=63 [rifampicin]), with all isolates carrying this substitution being daptomycin-resistant (Figure 4F). The H486Y substitution was identified at a reduced frequency compared to the S491F, (n=12 [rifaximin] and n=28 [rifampicin], respectively), with all isolates again being daptomycin-resistant. The G482D substitution was the third most commonly identified RpoB substitution (n=15 [rifaximin] and n=6 [rifampicin], respectively), with 19 isolates carrying this substitution being daptomycin-resistant. Other RpoB substitutions in addition to S491F, H486Y, and G482D were also identified. These substitutions included V135F, L471V, Q473L, and H486R, however all VREfm isolates carrying these substitutions were daptomycin-susceptible. Importantly, the proportions of each RpoB substitution observed in VREfm collected from the gastrointestinal tract of mice administered either rifaximin or rifampicin, closely matched the proportions of each mutation observed in our collection of human clinical VREfm isolates, with the S491F substitution most commonly identified, followed by H486Y, and then G482D. These observations in our mouse model strongly suggest that exposure to rifaximin is driving the *de novo* emergence of daptomycin resistance in colonising strains of *E. faecium* in humans.

### VREfm with daptomycin-resistant RpoB substitutions have changes in the cell membrane lipid profile and increased expression of the *prdRAB* regulon

To understand how amino acid substitutions in the ß subunit of the bacterial RNA polymerase (RpoB) cause resistance to daptomycin, a cell membrane active antibiotic, we utilised a multi-omics approach. We started by analysing the lipidomes of the three isogenic daptomycin-resistant RpoB mutants (G482D, H486Y, or S491F) compared to WT VREfm. We additionally included an isogenic RpoB mutant that did not confer resistance to daptomycin (Q473L; daptomycin MIC 2 mg L^-1^) created from the same WT VREfm isolate as a comparison. Principal-component analysis (PCA) of the detected lipids clearly separated the daptomycin-resistant RpoB mutants from the WT and daptomycin-susceptible Q473L strain, indicating distinct lipid profiles (Figure 5A). While the same classes of lipid species were observed in the WT and daptomycin-resistant RpoB mutants, there were significant reductions in anionic cardiolipins (CL) and phosphatidylglycerols (PG), as well as an increase in digalactosyldiacylglycerols (DGDG) and cationic lysyl-PGs (Lys-PG) in the RpoB mutants (Figure 5B-E). CL, PG, DGDG, and Lys-PG profiles returned to WT when each of the three RpoB mutations was reverted, demonstrating that the differences observed in the daptomycin-resistant strains were due to the RpoB substitutions (Supplementary Table 8).

**Figure 5.**
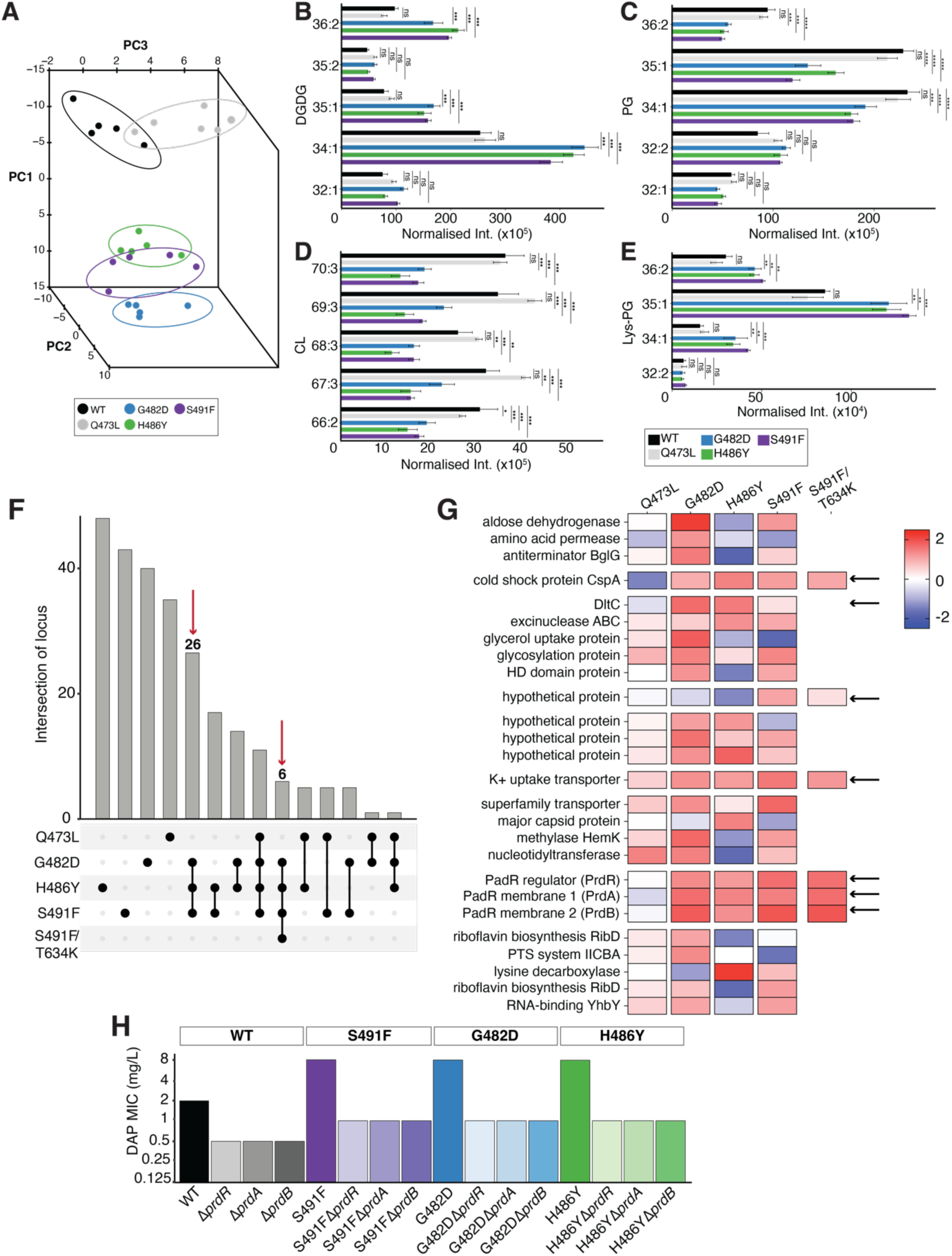
**A.** Principal-component analysis denoting segregation of the total lipid classes obtained for the WT and RpoB mutants. **B to E.** The lipid species differentially produced by the WT and RpoB mutants are showed as normalised abundance (intensity/total protein). Bars represent the median value and error bars represent the SEM (n=5). Data was analysed with a Two-way ANOVA (WT versus RpoB mutant) **F.** Intersection between RNAseq and proteomics analyses displayed as an UpSet plot (n=5 for RNAseq and n=5 for proteomics). RNA seq significance (FDR<0.05, log_2_FC > 1 or log_2_FC < -1) and proteomics significance (adjusted p-value<0.05, log_2_FC > 1 or log_2_FC < -1). **G.** Heatmaps displaying the 26 significantly, differentially expressed loci identified in both RNA seq and proteomics. The fold-change value for RNAseq is shown. **H.** Daptomycin susceptibility testing results for the WT and *rpoB* backgrounds with *prdR*, *prdA*, or *prdB* deleted (n=3). The MIC for each strain is shown without error bars since no variation between independent biological replicate was observed. ns not significant, \**P* < 0.05, \*\**P* < 0.01, \*\*\**P* < 0.001, \*\*\*\**P*<0.0001.

Given the central role of RpoB in transcription, we hypothesised the lipidome differences of the daptomycin-resistant RpoB mutants were caused by alterations in gene expression. We firstly modelled the RNA polymerase complex structure in *E. faecium* to estimate the changes imparted by Q473L, G482D, H486Y, and S491F substitutions on transcription at the protein level. All four substitutions were present at the rifamycin active site and in direct interaction with nucleic acids at the transcription replication fork, with predicted changes in stability or altered interactions with nucleic acid templates (Supplementary Figure 10). The highly prevalent S491F substitution defined changes to a bulkier hydrophobic side chain, with predicted mild reductions in protomer stability and affinities to rifampicin, other RNA polymerase subunits, and nucleic acids within the replication fork. Thereby suggesting that even at objectively low reductions in affinity, this mutation can directly impact the transcription rate, and the subsequent rate of gene expression. Conversely, the G482D substitution resulted in the introduction of a bulkier negatively charged side chain, which led to steric clashes and increased electrostatic potential of the RNA binding cleft. The G482D substitution was predicted to have the largest detrimental effect on protein stability, which may reduce the amount of active RNA polymerase and gene expression. Interestingly, it was also predicted to increase nucleic acid binding affinity of the mutant complex, further reducing RNA polymerase activity and processing. Finally, the daptomycin-resistant substitution H486Y and daptomycin-susceptible substitution Q473L were predicted to confer similar effects to the protein stability, and rifampicin and nucleic acid binding affinity. However, the H486Y substitution, but not the Q473 substitution, was predicted to increase RNA polymerase complex stability, reducing the dynamic flexibility required for enzyme activity, potentially leading to the changes in gene expression. Overall, the daptomycin-resistant substitutions S491F, H486Y, and G482D were all characterised by distinct interactions from the WT RNA polymerase while the daptomycin-susceptible substitution Q473L retained the original WT interactions.

Given the predicted changes in RNA polymerase transcriptional activity, we used a combination of RNA sequencing (RNAseq) and data independent acquisition (DIA) based proteomics and identified 26 loci that were significantly differentially expressed in both RNAseq and proteomics in all three daptomycin-resistant strains (G482D, H486Y, and S491F), but not in the daptomycin-susceptible strain (Q473L) or the WT VREfm (Figure 5F and Supplementary Table 9). Of note, there was no differences in expression (by transcriptomics or proteomics) of the *liaFSR*, *liaXYZ*, or *yycFG* (*walKR*) regulons or the cardiolipin synthase *(cls)* or division site tropomyosin (*divIVA*) genes, indicating the mechanism of RpoB-mediated daptomycin resistance was independent of previously described systems^10,11,21^. Since compensatory *rpoC* mutations can alter the kinetic parameters of the RNA polymerase enzyme, we hypothesised the number of dysregulated genes would decrease in the RpoBC double mutant (RpoB S491F and RpoC T634K) compared to the single RpoB S491F mutant, leaving genes likely associated with daptomycin resistance. Indeed, only six loci were differentially expressed (by RNAseq and proteomics) in the RpoBC mutant, compared with 44 loci identified in the single RpoB S491F mutant (Figure 5F-G). These included genes encoding a cold shock protein (CspA, [protein] AGS75480 or EFAU233_01583), a hypothetical protein of unknown function, a potassium uptake transporter (K+ transporter, [protein] AGS74117 or EFAU233_00176), and a DNA-binding transcriptional regulator of the PadR family in a putative operon with two hypothetical membrane proteins ([proteins] AGS74325, AGS74326, AGS74327 or EFAU233_00444, EFAU233_00445, EFAU233_00446). All six of these loci were significantly upregulated (in RNAseq and proteomics) in the three daptomycin-resistant RpoB mutants (G482D, H486Y, and S491F), as well as in the RpoBC double mutant.

To understand their potential role in daptomycin resistance, each of these six genes were deleted from the WT and RpoB S491F mutant. The *dltC* gene, linked to daptomycin resistance in *S. aureus*, was also included as it was differentially expressed in the G482D, H486Y, and S491F strains, but not in the RpoBC double mutant. Deletion of *cspA*, *dltC*, the hypothetical protein, or K+ uptake transporter did not alter daptomycin susceptibility in either the WT or RpoB S491F backgrounds. However, deletion of the PadR-family regulator (named here *prdR*, <u>P</u>henotypic <u>R</u>esistance to <u>D</u>aptomycin <u>R</u>egulator) or individual deletions of the hypothetical membrane proteins (named here *prdA* and *prdB*) increased daptomycin susceptibility by 4-fold in the WT or S491F backgrounds (Figure 5H; Supplementary Figure 11A). Similar 4-fold increases in daptomycin susceptibility were observed when these genes were deleted from the RpoB G482D or H486Y backgrounds (Figure 5H).

Clinical paired VREfm isolates representative of the G482D, H486Y, and S491F mutations were analysed by DIA proteomics to examine the abundance of PrdR, PrdA, and PrdB in the cell. In daptomycin-resistant clinical strains carrying the S491F (ST1421 and ST203), G482D (ST80), or H486Y (ST203) mutations, the production of PrdRAB was significantly (*P*<0.05) increased compared to daptomycin-susceptible (WT *rpoB* allele) strains of the same ST (Supplementary Figure 11B-C and Supplementary Table 9). The increased expression of PrdRAB in both genetically distinct clinical VREfm harbouring the G482D, H486Y, or S491F mutations and in isogenic strains, suggested the *prdRAB* operon was responsible for RpoB-mediated daptomycin resistance.

### Upregulation of the *prdRAB* operon leads to cell membrane remodelling and decreased daptomycin binding in daptomycin-resistant RpoB strains

We hypothesised the overexpression of the *prdRAB* operon in the RpoB mutants was leading to the observed changes in the cell membrane and subsequent resistance to daptomycin. Therefore, an overexpression vector with the *prdR* gene, along with an empty vector (EV) control, were introduced separately into the WT VREfm. Consistent with this, plasmid mediated overexpression of the *prdR* gene increased the WT daptomycin MIC 4-fold, to 8 mg L^-1^. Proteomic analysis (WT EV versus WT*^prdR^*) confirmed the *prdR* regulator was expressed to a similar level in the WT*^prdR^* strain as in the RpoB mutants (2.5-fold and 2.2-fold in S491F, respectively) and the *prdR* regulator specifically controlled the expression of the *prdAB* membrane proteins (Supplementary Figure 12A and Supplementary Table 9). PCA of the WT, WT EV, WT*^prdR^*, and RpoB S491F lipidomes supported the overexpression of the *prdR* gene driving the changes in the lipid species, with the WT*^prdR^* and RpoB S491F lipidomes clustering separately than the WT and WT EV lipidomes (Figure 6A and Supplementary Table 8).

**Figure 6.**
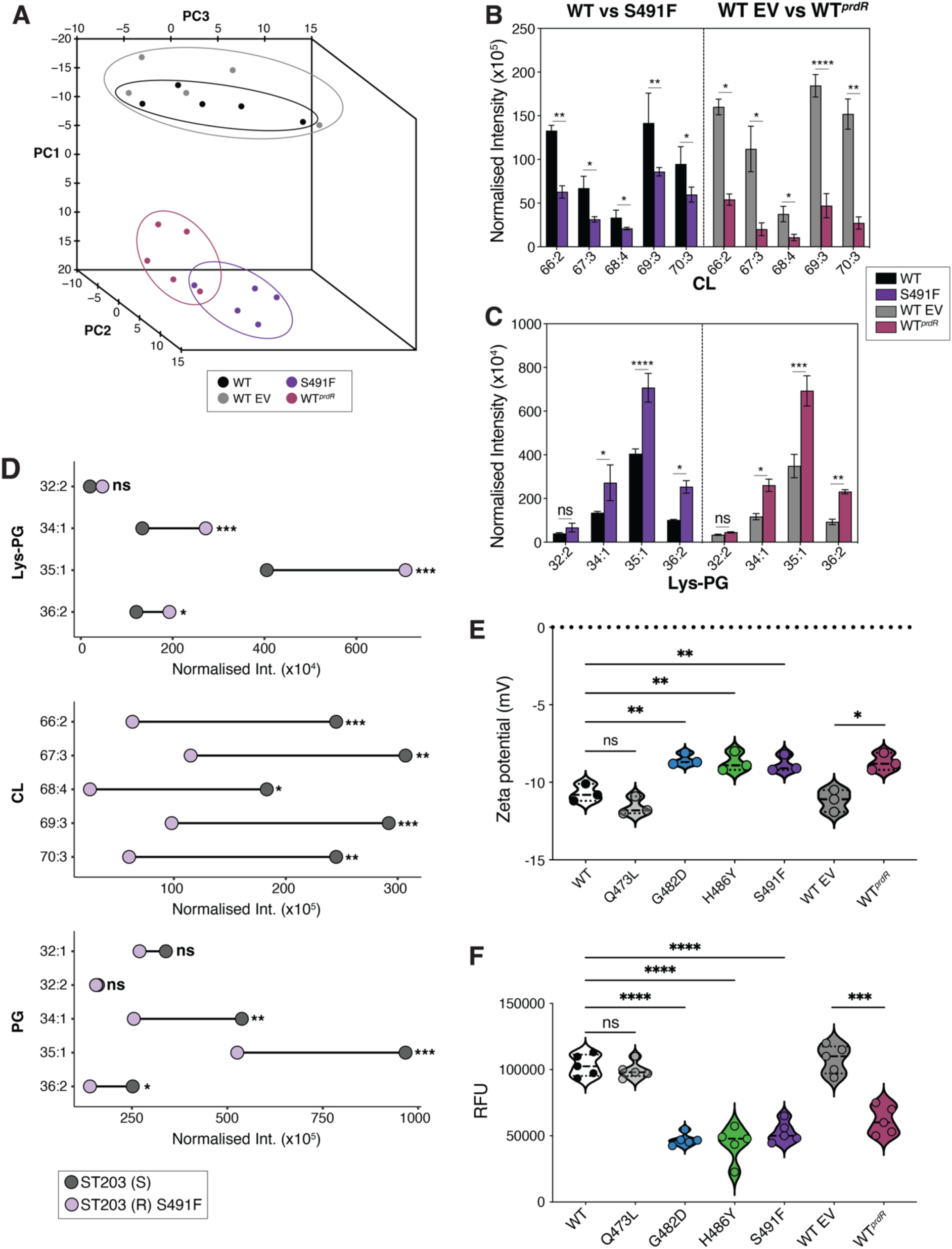
**A.** Principal-component analysis denoting segregation of the total lipid classes obtained for the WT, S491F mutant, WT empty vector (EV), and WT*^prdR^*. **B and C.** The lipid species differentially produced by the WT, S491F mutant, WT EV, and WT*^prdR^* are showed as normalised abundance (intensity/total protein). Bars represent the median value and error bars represent the SEM (n=5). Data was analysed with a Two-way ANOVA (WT versus RpoB mutant and WT EV versus WT*^prdR^*). **D.** Lipid species differentially produced by the ST203 RpoB S491F clinical strain pair, showed as normalised abundance (intensity/total protein). The dots represent the median value (n=5). Data was analysed with a Two-way ANOVA (ST203 DAP-S versus ST203 DAP-R). Lipid species with a significant difference are denoted. **E.** Zeta potential (measured in mV). Points represent each biological replicate (n=3) and lines represent the median and interquartile range. Data was analysed with a Two-way ANOVA (WT versus RpoB mutant and WT EV versus WT*^prdR^*). **F.** Binding of BoDIPY-DAP, represented as relative fluorescence units (RFU). Points represent each biological replicate (n=5) and lines represent the median and interquartile range. Data was analysed with a Two-way ANOVA (WT versus RpoB mutant and WT EV versus WT*^prdR^*). ns not significant, \**P* < 0.05, \*\**P* < 0.01, \*\*\**P* < 0.001, \*\*\*\**P*<0.0001.

Reductions in the same anionic phospholipids (CL and PG) and increases in cationic Lys-PG species were observed in the WT*^prdR^* strain to similar levels seen in the RpoB S491F mutant (Figure 6B-C and Supplementary Figure 12B). Lipidomic analyses of the previously described paired clinical isolates demonstrated the daptomycin-resistant clinical strains carrying RpoB mutations had similar significant differences in charged phospholipids (decreases in CL and PG, increases in Lys-PG), compared to daptomycin-susceptible strains containing the WT *rpoB* allele (Figure 6D and Supplementary Table 8). These data indicated that overexpression of the *prdRAB* operon in daptomycin-resistant VREfm carrying RpoB mutations leads to changes in the abundance of charged lipid species in the cell membrane.

Given the decreases in anionic phospholipids (CL and PG) and increases in cationic phospholipids (Lys-PG), we tested if strains harbouring RpoB mutations harbour differences in overall cell membrane charge and daptomycin binding. As we predicted, both the daptomycin-resistant isogenic and clinical RpoB mutants (G482D, H486Y, and S491F) had a significantly reduced negative charge associated with the cell membrane compared to their paired isolate (Figure 6E and Supplementary Figure 13A), respectively, and consequently bound less fluorescent daptomycin (Figure 6F and Supplementary Figure 13C). Complementation of the RpoB mutations reversed cell membrane charge and daptomycin binding to WT levels (Supplementary Figure 13B and D). No significant difference was observed in membrane charge or daptomycin binding for the daptomycin-susceptible Q473L strain (Figure 6E-F). Further, the overexpression of the *prdR* gene resulted in a significantly reduced negative charge across the cell membrane compared to the WT EV strain, with a similar charge to the daptomycin-resistant RpoB mutants (Figure 6F). Additionally, binding of daptomycin was significantly decreased compared to the WT EV strain (Figure 6F). These experiments show that the increased expression of the *prdRAB* operon in VREfm carrying daptomycin resistance associated RpoB mutations leads to changes in the abundance of charged phospholipids and a decrease in the cell surface negative charge, which in turn reduces daptomycin binding to the cell membrane.

## Discussion

Here we show that specific amino acid substitutions within the RRDR of RpoB (G482D, H486Y, and S491F) lead to clinical daptomycin resistance in *E. faecium*. These mutations are globally distributed with prevalence equal to other well-characterised amino acid substitutions within LiaFSR/LiaXYZ and Cls, which have been considered the dominant mechanisms of daptomycin resistance in VREfm^21,38,39^. Although these findings are reminiscent of previous studies in *S. aureus,* which have suggested that *rpoB* mutations can lead to reduced daptomycin susceptibility, there are some important distinctions^40–42^. In *S. aureus, rpoB* mutations have not been reported to result in clinical daptomycin resistance (defined as MIC≥ 2 mg L^-1^), as seen here in VREfm - rather they are associated with subtle changes in daptomycin susceptibility. There are no reports in the literature of *rpoB* mutations leading to daptomycin resistance in clinical *S. aureus* isolates, suggesting that *rpoB* mutations may be of limited clinical importance to *S. aureus* daptomycin resistance. This contrasts with VREfm, where *rpoB* mutations appear to be a major mechanism of daptomycin resistance. Through our multi-omic approach we have defined the molecular mechanism by which *rpoB* mutations lead to changes in daptomycin susceptibility in VREfm. Our analyses show that G482D, H486Y, and S491F RpoB substitutions mediate daptomycin resistance via a conserved mechanism that is independent of previously described systems (Lia operons or Cls). Each of these RpoB substitutions caused similar transcriptional reprograming in VREfm, with dysregulation of a previously uncharacterised genetic locus found to be responsible for daptomycin resistance. The locus, that we have named the <u>P</u>henotypic <u>R</u>esistance to <u>D</u>aptomycin (*prd*) operon consists of a transcriptional regulator (PrdR) and two putative membrane proteins (PrdAB). Upregulation of the *prdRAB* operon leads to cell membrane remodelling, with decreased levels of anionic phospholipids (PG and CL) and increased levels of cationic phospholipids (Lys-PG). This ultimately decreases the negative cell surface charge and reduces daptomycin binding, which renders VREfm resistant to daptomycin. Notably, there was no dysregulation of the *prdRAB* operon in an isogenic VREfm strain carrying an RpoB substitution (Q473L) that did not confer daptomycin resistance and no membrane changes relative to the daptomycin-susceptible WT were observed. The G482D, H486Y, and S491F RpoB substitutions studied here should therefore be considered clinically relevant and major mediators of daptomycin resistance in VREfm.

The S491F RpoB substitution in particular is common within the global *E. faecium* population, being present in at least three phylogenetically distinct lineages currently circulating within healthcare systems in Australia, the United Kingdom, the United States, and across Europe. These lineages have spread over global geographic scales and persisted for at least 15 years, which is of clinical concern since it suggests that they might eventually compromise the therapeutic value of daptomycin for treating VREfm infections.

Exposure to rifamycin antimicrobials is a well-documented driver in the emergence of bacterial strains carrying substitutions in RpoB, with mutations in this gene often leading to high level rifamycin resistance^24^. In keeping with this, VREfm isolates carrying the G482D, H486Y, and S491F RpoB substitutions are resistant to rifamycin antimicrobials in addition to daptomycin, suggesting that rifamycin use might be an important driver in the emergence of daptomycin-resistant VREfm. In support of this, our data suggests the clinical use of rifaximin is likely responsible for driving VREfm isolates harbouring mutations within *rpoB* that are cross-resistant to rifamycins and daptomycin. Three lines of evidence support this hypothesis: (i) Bayesian phylodynamic analyses show the emergence of phylogenetically distinct VREfm lineages carrying the S491F substitution is temporally linked with the clinical approval of rifaximin in the early 2000s, with each lineage subsequently undergoing population expansion following FDA approval in 2010 of rifaximin as a prophylactic for the prevention of hepatic encephalopathy in chronic liver disease patients, (ii) two independent retrospective patient cohort studies demonstrate that recent rifaximin exposure was predictive of carriage of daptomycin-resistance RpoB substitutions, and (iii) animal experiments demonstrate the administration of rifaximin to mice colonised with VREfm led to the *de novo* emergence of VREfm strains within the gastrointestinal tract that carried substitutions within RRDR of RpoB and were resistant to rifamycins and cross-resistant to daptomycin.

Although both rifaximin and rifampicin could drive the *de novo* emergence of daptomycin-resistant VREfm in the mice experiments of our study, the data overall do not support a prominent role for rifampicin in the emergence of daptomycin-resistant VREfm. This hypothesis is based on a lack of temporal signal between the emergence of daptomycin-resistant VREfm carrying RpoB substitutions (circa 2006) and the clinical introduction of rifampicin which occurred in the early 1970s, as well as the observation that the vast majority of daptomycin-resistant VREfm were only isolated from patients who received rifaximin in our clinical analyses (and not from the control group). Why rifaximin appears to be a driver of daptomycin resistance in *E. faecium* while rifampicin does not, is not immediately clear, but may relate to the patient cohorts that rifampicin and rifaximin are primarily used to treat. Rifaximin is used in patients with chronic liver disease who are recognised as one of the highest risk groups for gastrointestinal VREfm carriage, while rifampicin and other rifamycins are used in broader cohorts that are not frequently colonised with *E. faecium*. Our hypothesis is that patients with chronic liver disease receiving rifaximin are a major source for the emergence and subsequent spread of daptomycin-resistant VREfm carrying *rpoB* mutations. Our analysis of the HSCT patient cohort demonstrates that this phenomenon is not restricted to liver disease patients and is more generalisable, such that any patient colonised with VREfm and receiving rifaximin is at risk of *E. faecium* carrying *rpoB* mutations that are daptomycin-resistant emerging within the gut. There are some important clinical implications from this. First, our results suggest that daptomycin should not be used for empiric therapy of invasive VREfm infections in patients who are receiving rifaximin, or have recently received rifaximin, given the significantly higher risk of daptomycin resistance. Secondly, to preserve the use of daptomycin for VREfm, consideration should be given to maintaining isolation precautions in hospitals for patients receiving rifaximin who are colonised with VREfm, avoiding cohorting with other VREfm colonised patients where possible. Thirdly, although effective for prophylaxis against hepatic encephalopathy, consideration should be given to keeping rifaximin as a second-line option behind other therapies for this indication, and the use of rifaximin for prophylaxis following HSCT reconsidered, given the propensity to induce mutations in *rpoB* and subsequent daptomycin resistance.

There is a coordinated global effort to preserve the use of last-resort antimicrobials through strict stewardship protocols that try to limit the use of these critically important medicines. The rationale for this strategy assumes that restricted antibiotic usage equals less opportunity for pathogens to develop resistance. Here we show that this assumption may be flawed, since exposure to prophylactic rifaximin can lead to the emergence of daptomycin-resistant VREfm in the absence of daptomycin exposure. These data highlight that unanticipated antimicrobial cross-resistance can readily arise following the implementation of new antimicrobial regimes, even when they are perceived to be “low-risk”. Careful consideration should therefore be given to the potential impact of prophylactic antibiotics on antimicrobial stewardship practices.

We have identified a previously unrecognised, globally important mechanism of last-resort antibiotic resistance in VREfm and show that prophylactic rifaximin use, particularly in chronic liver disease patients, is a cause of this resistance. These findings demonstrate the ease with which new antibiotic treatment regimens can drive the emergence of multidrug-resistant pathogens and highlight the impact that unanticipated antibiotic cross-resistance can have on antibiotic stewardship efforts designed to preserve the use of last-resort antibiotics. We advocate for the judicious use of all antibiotics.

## Methods

### Media and reagents

*E. faecium* was routinely cultured at 37°C in brain heart infusion (BHI) broth (Becton Dickson) or BHI agar (BHIA), BHI solidified with 1.5% agar (Becton Dickson). For electroporation, *E. faecium* was cultured in BHI supplemented with 3% glycine and 200 mM sucrose (pH 7.0). *E. coli* was cultured in Luria broth (LB). Broth microdilution (BMD) MICs were performed in cation-adjusted Mueller Hinton with TES broth (CAMHBT) (Thermo Fisher). A concentration of 10 mg L^-1^ chloramphenicol (Sigma Aldrich) was used for plasmid selection in *E. faecium* and *E. coli*. The following antibiotics were used at variable concentrations for susceptibility testing: rifampicin (Sigma Aldrich), rifaximin (Sigma Aldrich), and daptomycin (Cubicin). Oligonucleotides were purchased from Integrated DNA Technologies and are listed in Supplementary Table 11. Plasmids were purified with Monarch Plasmid Miniprep Kit (NEB). PCR products and gel extractions were purified using Monarch DNA Gel Extraction Kit (NEB). Genomic DNA was purified using the Monarch Genomic DNA Purification Kit (NEB). Phusion and Phire DNA polymerase was purchased from NEB.

### Bacterial isolates

Bacterial strains used in this study are listed in Supplementary Table 10. Australian bacterial strains were collected across three data projects in the Microbiological Diagnostic Unit Public Health Laboratory (MDU PHL). Two unbiased cross-sectional surveys of VREfm were conducted between 10 November and 9 December 2015 (n=331)^32^ and between 1 November and 30 November 2018 (n=323) in the State of Victoria (referred to as the 2015 and 2018 Snapshot). During this period, all VREfm-positive isolates (including screening and clinical samples) collected by laboratories across the state were sent to the MDU PHL. In addition, this project included *vanA*-VREfm collected from the “Controlling Superbugs” study^19,20^, a 15-month (April-June 2017 and October 2017-2018) prospective study including eight hospital sites across four hospital networks, resulting in 346 VREfm isolates (308 patients) sent for WGS at MDU PHL. The VREfm were isolated from patient samples (including screening and clinical samples) routinely collected from hospital inpatients. For the ‘historical *vanA*-VREfm,’ every *vanA* isolate collected within MDU PHL was included. This resulted in an additional 229 isolates, sampled between 2003 and 2014.

For publicly available isolates, our aim was to capture the diversity of *E. faecium* circulating globally by including isolates that formed part of several key studies involving hospital-associated VREfm (as of January 2021). To be included, isolates needed to have short-read data available, with geographic location (by country), year of collection, and source (human or animal). Reads were only included if they had a sequencing depth of >50x. To capture the diversity of VREfm circulating in the United States, isolates from human sources were downloaded from the PathoSystems Resource Integration Center^43^. All isolates were confirmed to be *E. faecium* with the Kraken2 database (v.2.1.2)^44^. The final number of international isolates comprised those from Africa (n=8), Asia (n=25), Europe (n=2941), North America (n=424), and South America (n=78) (Supplementary Table 10).

### Antibiotic susceptibility testing

Daptomycin susceptibility testing was performed using the BMD MIC method according to CLSI guidelines. In a 96-well plate, a two-fold dilution series (from 32 to 0.5 mg L^-1^) of daptomycin was made in 100 μL volumes of CAMHBT, additionally supplemented with 50 mg L^-1^ Ca^2+^. An inoculum of 100 μL *E. faecium* broth culture adjusted to 1 x 10^6^ CFU mL^-1^ in CAMHBT was then added to each well. After 24 hours incubation, the MIC was defined as the lowest antimicrobial concentration that inhibited visible growth. All assays were performed in biological triplicate, with the median MIC reported. In accordance with recent guidelines^45^, isolates with a daptomycin MIC ≥ 8 mg L^-1^ were considered to be daptomycin-resistant. A daptomycin-sensitive strain (AUS0085)^46^ and a daptomycin-resistant strain (DMG1700661)^16^ were used as a control.

Rifampicin susceptibility testing was performed using the BMD method in CAMHBT. High-level rifampicin resistance was defined with a MIC > 32 mg L^-1^. All susceptibility testing was performed in triplicate.

### Whole-genome sequencing

Genomic DNA was extracted from a single colony using a JANUS automated workstation (PerkinElmer) and Chemagic magnetic bead technology (PerkinElmer). Genomic DNA libraries were prepared using the Nextera XT kit according to manufacturer’s instructions (Illumina, San Diego, CA, USA). Whole-genome sequencing was performed using Illumina NextSeq platform, generating 150 bp paired-end reads. The short reads of isolates sequenced at MDU-PHL are available on the NCBI Sequence Read Archive [BioProjects PRJNA565795 (Controlling Superbugs), PRJNA433676 (2015 Snapshot) and PRJNA856406 (2018 Snapshot), and PRJNA856406 (historical *vanA* isolates)].

### Phylogenetic analysis

De novo assemblies of the genomes were constructed using Spades^47^ (v3.13). In silico MLST was determined using the program mlst with the efaecium database^48^ (https://github.com/tseemann/mlst). The 1000 Australian genomes as well as the 4,705 Australian and international VREfm were mapped to the reference *E. faecium* genome AUS0085 isolated from a human bacteraemia infection in Victoria, Australia (NCBI accession: CP006620)^46^ using snippy (https://github.com/tseemann/snippy) (v4.4.5), applying a minfrac value of 10 and mincov value of 0.9. This reference was selected as it was a publicly available complete genome collected locally and daptomycin-susceptible. A maximum likelihood phylogenetic tree was inferred using IQ-TREE (v2.1.2)^49^ with a general time-reversible (GTR + G4) substitution model, including invariable sites as a constant pattern and 1000 bootstrap replicates. Recombination masking was not performed for species maximum likelihood trees due to the small size of the resulting core alignment. All trees were mid-point rooted and visualised in R (v4.0.3, https://www.r-project.org/) using phangorn^50^ (v2.5.5), ape^51^ (v5.4), ggtree^52^ (v2.3.4), and ggplot (v3.3.2).

The genome assemblies of all isolates were screened for acquired antimicrobial resistance determinants using abriTAMR (https://github.com/MDU-PHL/abritamr)^53^.

### Genome-wide association study of daptomycin resistance

A GWAS approach was applied to identify genetic variants of daptomycin resistance in *E. faecium*. A genotype matrix of SNPs was constructed and used as input to homoplasyFinder^54^ (v0.0.0.9) to determine the consistency index at each locus and kept mutations that had an index of ≤0.5 (indicating at least two independent acquisitions across the phylogeny). We then ran GWAS using daptomycin resistance as a binary trait, where isolates were categorised as resistant if their daptomycin MIC was ≥8 mg L^-1^. To correct for population structure, we used the factored spectrally transformed linear mixed models (FaST-LMM) implemented in pyseer^55^ (v.1.3.6), which computes a kinship matrix based on the core genome SNPs as a random effect. *P* values were corrected for multiple hypothesis testing using the Bonferroni correction method.

### Competition assay

For competition assays, overnight cultures of WT and corresponding RpoB S491F or RpoB S491F/RpoC T634K mutants were diluted to an OD600 of 0.5 in BHI and equal volumes added to an overnight culture. Serial dilutions of each co-culture were performed at times 0 and 24 hours on BHIA. Colonies were then replica plated onto BHIA and BHIA rifampicin 20 µg mL^-1^ to determine the proportion of WT to mutant.

### Core genome MLST (cgMLST) and clustering

cgMLST alleles for each isolate was defined using the public *E. faecium* cgMLST scheme^56^ and chewBBACA (v2.0.16)^57^, implemented locally in the COREugate pipeline (v2.0.4) (https://github.com/kristyhoran/Coreugate). The pipeline determines the alleles of each core gene for every isolate as defined by the specific pathogen scheme. The *E. faecium* cgMLST scheme contains 1,423 genes. The number of allelic differences between each isolate within this core set of genes is then determined. The cgMLST clusters were determined using single linkage clustering and a pairwise allelic difference threshold of ≤250. This threshold was chosen since it maximised diversity within clusters, to improve temporal sampling depth, while still clustering based on maximum-likelihood tree structure.

### Phylodynamic analyses of the emergence of the S491F RpoB mutation in VREfm lineages

To investigate the emergence of the S491F mutation in RpoB in three different lineages, as defined with cgMLST, we undertook further analysis on these clusters/lineages. From the species-level maximum-likelihood tree (Figure 2C), three lineages/clusters were identifiable by cgMLST due to their size (n>50) and presence of the S491F mutation. The three clusters were analysed independently, such that individual core-genome SNP alignments were generated, since this increased the length of the core alignment and number of sites considered. Snippy (https://github.com/tseemann/snippy) (v4.4.5) was used to generate the alignments for each cluster to the corresponding reference genome (AUSMDU00004024/CP027517.1 for Cluster 1, AUSMDU00004055/CP027506.1 for Cluster 2, and AUSMDU00004142/CP027501.1 for Cluster 3). Each core alignment used a within ‘cluster reference’ (complete genome of the same cluster) to maximise core-SNP alignment length.

The reference for each cluster was chosen since they were a locally-collected, closed genome. Recombination was removed from the final alignment using Gubbins^58^ (v.2.4.1) to ensure modelling was only informed by SNPs with tree-like evolution within the core genome. Maximum-likelihood trees for each of the three clusters were inferred from the core-SNP alignments [Cluster 1: (n=219 taxa) 329 SNPs; Cluster 2: (n=85 taxa) 541 SNPs; Cluster 3: (n=68 taxa) 764 SNPs] with IQ-tree (v2.1.2)^59^ with a general time-reversible (GTR+G4) substitution model, including invariable sites as a constant pattern. Phylogenetic uncertainty was determined through 1000 nonparametric bootstrap replicates.

To investigate temporal signal in the three clusters of VREfm genomes, we first used TempEst^60^ (v1.5). A root-to-tip regression analysis was performed on the root-to-tip branch distances within the three, cluster maximum-likelihood phylogenies as a function of year of collection, with the position of the root optimised according to the heuristic residual mean squared method.

The frequency of the emergence of the *rpoB* mutation in VREfm was inferred using a discrete trait model implemented in BEAST^61^ (v1.10.4). Under this model the SNP alignments are used to infer the evolutionary process (i.e. phylogenetic tree, time, and nucleotide substitution model parameters) for the three clusters. The alignments all shared the HKY substitution model and a constant-size coalescent population prior^26^. To avoid ascertainment bias due to using a SNP alignment, the number of constant sites were considered for the likelihood calculations. The molecular clock was a relaxed clock with an underlying lognormal distribution. The molecular clock was calibrated using isolation dates for each genome by year of collection and the mean clock rate is shared between all three alignments, but the model allows for the individual alignments to have different standard deviations of the lognormal distribution and also different branch rates. The mean molecular clock rate requires an explicit prior distribution, for which we used a Gamma distribution and a 0.95 quantile range of 4.9×10^-6^ and 1.1×10^-4^ substitutions/site/year. This informative prior means that it acts as an additional source of molecular clock calibration.

The presence or absence of the S491F mutation in *rpoB* was used as a binary trait^62,63^. The trait model was shared between the three alignments, with the different Markov jumps and rewards (i.e. changes of trait state and time spent in each state, respectively) recorded for each of the three alignments. The posterior distribution of model parameters was sampled using a Markov chain Monte Carlo of 100,000,000 iterations, sampling every 100,000 iterations. Two independent runs were run for the models. We assessed sufficient sampling from the stationary distributions by verifying the effective sample size of key parameters was around or above 200. The final maximum-clade credibility trees were visualised in R (v4.0.3, https://www.r-project.org/) using ggtree^52^ (v2.3.4). The Markov jumps for the *rpoB* trait for each alignment were visualised in R (v4.0.3, https://www.r-project.org/).

### Construction of isogenic mutants using allelic exchange and of pRAB11*^prdR^*

The *rpoC*^T634K^, *rpoB*^G482D^, *rpoB*^H486Y^, *rpoB*^S491F^, *rpoB*^Q473L^, ABC transporter (I274S), permease protein (G71S), or mannitol dehydrogenase (V288L) mutations were recombined into the chromosomal copy of each gene in ST796 VREfm (Ef_aus0233) by allelic exchange. Deletions of the CpsA, K+ transporter, hypothetical protein, DltC, or PrdRAB were also completed using allelic exchange. The region encompassing each gene was amplified by SOE-PCR and recombined into pIMAY-Z^64^ by the seamless ligation cloning extract (SLiCE)^65^ method and transformed into *E. coli* IM08B^64^. The construct was transformed into electrocompetent VREfm^65^, with allelic exchange performed as described previously^66^. Reversion of *rpoB*^G482D^, *rpoB*^H486Y^, or *rpoB*^S491F^ mutations were completed using allelic exchange with a construct containing the respective wild-type allele. To construct a vector containing *prdR*, the vector pRAB11 was used. The *prdR* gene was amplified using Aus0233 genomic DNA. The *prdR* product was gel extracted, SLiCE cloned into amplified pRAB11, and transformed into IMO8B, yielding pRAB11:*prdR*. The plasmid and empty vector was then electroporated into Aus0233. Genome sequencing and analysis of all mutants was conducted as described, with resulting reads mapped to the Ef_aus0233 reference genome and mutations identified using Snippy (https://github.com/tseemann/snippy) (v4.4.5).

### VREfm *in vivo* gastrointestinal colonisation experiments

Female C57BL/6 mice at 6-8 weeks of age were purchased from WEHI and maintained in a specific-pathogen-free facility at the Peter Doherty Institute for Infection and Immunity. All animal handling and procedures were performed in a biosafety class 2 cabinet. Animal procedures were performed in compliance with the University of Melbourne guidelines and approved by the University’s Animal Ethics Committee.

The dose for each antibiotic was calculated using the FDA human conversion formula to ensure each mouse was given a human-equivalent dose^67^. To establish gastrointestinal colonisation of VREfm, mice were administered ceftriaxone (410 mg kg^-1^ day^-1^; AFT Pharmaceuticals) via subcutaneous injection once daily for 4 days, followed by an antibiotic wean period of 24 hours. Mice were then inoculated with 10^6^ VREfm in 100 μl PBS by oral gavage. Three days after VREfm inoculation, single-housed mice were administered either rifaximin (113 mg kg^-1^ administered twice daily; Sigma Aldrich), rifampicin (123 mg kg^-1^ administered once day; Sigma Aldrich), or vehicle (Corn oil with 10% DMSO) via oral gavage; or daptomycin (50 mg kg^-1^ administered once daily; Cubicin) via subcutaneous injection [this results in similar exposure (AUC) to that observed in humans receiving 8 mg kg^-1^ of intravenous daptomycin^68^]. The above antibiotic dosing protocol was followed for 7 days. Faecal samples were collected at specific time points throughout the experiment to determine VREfm gut colonisation and for downstream rifamycin and daptomycin resistance analysis. Faecal samples were resuspended in PBS to a normalised concentration (100 mg ml^-1^). Serial dilutions were performed, and samples were plated onto Brilliance VRE agar (Thermo Fisher) for VREfm CFU enumeration.

For rifamycin and daptomycin analysis, VREfm colonies (n=50 per mouse) from the Brilliance VRE agar plates were replica plated onto BHIA with and without rifampicin 20 µg mL^-1^ to determine the proportion of rifampicin-resistant VREfm in each mouse. Fifty colonies per mouse were then screened for daptomycin resistance, with a single colony being resuspended in PBS, then diluted 1/100 into CAMHBT containing 50 mg L^-1^ Ca^2+^, and 1/100 in CAMHBT containing 50 mg L^-1^ Ca^2+^ and 8 mg L^-1^ daptomycin. All suspected daptomycin-resistant colonies were confirmed using a daptomycin BMD MIC as before.

To determine which mutations were present in the rifamycin-resistant isolates, a random selection of 300 colonies, 150 from rifaximin treated mice and 150 from rifampicin treated mice (50 pre and 100 post for each treatment), were sampled for WGS as described above.

### Analysis of patients receiving rifaximin for hepatic encephalopathy prophylaxis

To examine the potential association between rifaximin exposure and daptomycin-resistant VREfm, we analysed VREfm collected between 2014 and 2022 from a single quaternary hospital institution in Melbourne. 225 patients were assessed for previous exposure to rifaximin, which was defined as at least a single dose administered prior to the collection date for the VREfm isolate, and grouped into “Rifaximin-exposed” and unexposed “Control” groups. Only a single isolate was selected at random for testing and analysis from patients who had multiple samples with VREfm isolates. The VREfm isolates underwent WGS and daptomycin and rifampicin susceptibility testing as before. Patients with VREfm isolates that were assessed as genetically clustered with other VREfm isolates in the cohort and likely represented direct transmission were excluded. Genetic clustering was defined using an international standard SNP cut-off (7 SNPs)^69,70^ using a split Kmer (k=15) analysis (SKA) (https://github.com/simonrharris/SKA) (v.1.0), a reference-free pairwise method that compares the entire genome (unlike traditional core-genome based comparisons). Medical records from patients were reviewed for comorbidity and antibiotic prescribing data. Potential associations were assessed through univariate analysis using Pearson’s chi-squared test or Fisher’s exact test for categorical data, and the Student’s *t*-test (parametric) or Wilcoxon Rank Sum test (non-parametric) for continuous data. To determine predictors of daptomycin-resistant VREfm, multivariable logistic regression with backward stepwise elimination procedure was used, excluding variables with *P*>0.10 and re-including variables with *P*<0.05. Exposure to rifampicin and daptomycin were forced into the models as variables *a priori*. A *P* value of <0.05 was considered statistically significant. Cases with missing data (e.g. incomplete medical records data due to inter-hospital transfer) were excluded. Several sensitivity analyses were also performed to assess for independent associations after excluding potential confounders, including analysis with exclusion of variables with <10 outcomes, assessing associations with antimicrobial exposure separately from demographic and comorbidity data, and modifying the rifaximin exposure variable to include a) any prior exposure to rifaximin (including both recent and distant exposure), and b) any prior exposure to rifamycin antibiotics (including rifampicin, rifabutin and rifaximin). The genomic relationships of VREfm isolates were visualised in a maximum-likelihood phylogenetic tree as before, using a core-SNP alignment of 12886 sites. The mutations in RpoB were determined using snippy (https://github.com/tseemann/snippy) (v4.4.5) as described.

### Analysis of patients receiving rifaximin for haematopoietic stem cell transplantation prophylaxis

To examine the potential association between rifaximin exposure and the presence of daptomycin associated *rpoB* substitutions in VREfm independent of underlying chronic liver disease, we analysed isolates collected from patients undergoing haematopoietic stem cell transplantation (HSCT) from a hospital institution in Regensburg, Germany. In this cohort, rifaximin is used for gut decontamination to reduce risk of gastrointestinal graft-versus-host-disease, while liver cirrhosis is a contraindication to HSCT. There were 68 patients initially assessed for recent exposure to rifaximin, which was defined as at least a single dose administered within 90 days prior to the isolate collection date. Only a single isolate was retained for testing and analysis from patients who had multiple samples. In this instance, the isolate included was randomised. The isolates underwent WGS using Ion Torrent Next-Generation sequencing technology (Thermo Fisher) and nanopore (Oxford Nanopore). Patients with isolates that were assessed as genetically clustered with other isolates in the cohort and likely represented direct transmission were excluded (as above). Statistical and phylogenetic analyses were undertaken as described above.

### Ultrahigh-performance liquid chromatography (UHPLC) and mass spectrometry (MS) analyses

Cultures of VREfm (n=5) were grown to mid-exponential phase (OD600 = 0.6) and washed in PBS. The protein content for each sample was measured with a Pierce BCA Protein Assay Kit (Thermo Fischer) and normalised to 100 μg. Cells were lysed using Bertin Precellys 24 homogenizer set at 6000 rpm for 40 seconds and lipids were subjected to monophasic extraction as described previously^72^. Lipidomic samples were analysed by UHPLC coupled to tandem mass spectrometry (MS/MS) employing a Vanquish UHPLC linked to an Orbitrap Fusion Lumos mass spectrometer (Thermo Fisher Scientific, San Jose, CA, USA), with separate runs in positive and negative ion polarities. Solvent A [60:40 (v:v) acetonitrile/water with 5 mM medronic acid and 10 mM ammonium acetate] and solvent B [90:10 (v:v) isopropanol:acetonitrile with 10 mM ammonium acetate]. 10 µL of each sample was injected into an Acquity UPLC HSS T3 C18 column (1 x 150 mm, 1.8 µm; Waters, Milford, MA, USA) at 50 °C at a flow rate of 60 μL/min for 3 min using 3% solvent B. During separation, the percentage of solvent B was increased from 3% to 70% in 5 min and from 70% to 99% in 16 min. Subsequently, the percentage of solvent B was maintained at 99% for 3 min. Finally, the percentage of solvent B was decreased to 3% in 0.1 min and maintained for 3.9 min.

All MS experiments were performed using an electrospray ionization source. The spray voltages were 3.5 kV in positive ionisation-mode and 3.0 kV in negative ionisation-mode. In both polarities, the flow rates of sheath, auxiliary and sweep gases were 25 and 5 and 0 arbitrary unit(s), respectively. The ion transfer tube and vaporizer temperatures were maintained at 300 °C and 150 °C, respectively, and the ion funnel RF level was set at 50%. In the positive ionisation-mode from 3 to 24 min, top speed data-dependent scan with a cycle time of 1 s was used. Within each cycle, a full-scan MS-spectra were acquired firstly in the Orbitrap at a mass resolving power of 120,000 (at m/z 200) across an m/z range of 300–2000 using quadrupole isolation, an automatic gain control (AGC) target of 4e5 and a maximum injection time of 50 milliseconds, followed by higher-energy collisional dissociation (HCD)-MS/MS at a mass resolving power of 15,000 (at m/z 200), a normalised collision energy (NCE) of 27% at positive mode and 30% at negative mode, an m/z isolation window of 1, a maximum injection time of 35 milliseconds and an AGC target of 5e4.

### Identification and quantification of lipids and statistical analysis

LC-MS/MS data was searched through MS Dial 4.90. The mass accuracy settings were 0.005 Da and 0.025 Da for MS1 and MS2. The minimum peak height was 50000 and mass slice width was 0.05 Da. The identification score cut off was 80%. In positive ionisation mode, [M+H]+, [M+NH4]+ and [M+H-H2O]+ were selected as ion forms. In negative ionisation mode, [M-H]-and [M+CH3COO]-were selected as ion forms. All lipid classes available were selected for the search. PC, Lys-PC, DG, TG, CE, SM were identified and quantified from positive ionisation mode while PE, LPE, PS, LPS, PG, LPG, PI, LPI, PA, LPA, Cer, CL were identified and quantified in negative ionisation mode. The retention time tolerance for alignment was 0.1 min. Lipids with maximum intensity less than 5-fold of average intensity in blank were removed. All other settings were default. All lipid LC-MS features were manually inspected and re-integrated when needed. These four types of lipids, 1) lipids with only sum composition except SM, 2) lipid identification due to peak tailing, 3) retention time outliner within each lipid class, 4) LPA and PA artefacts generated by in-source fragmentation of LPS and PS were also removed. The shorthand notation used for lipid classification and structural representation follows the nomenclature proposed previously^73^. Relative quantification of the lipid species was achieved using the mass spectrometry intensity of each lipid ion at apex of LC peak and normalized to the protein quantity in each sample.

### RNA-seq transcriptomics analysis

Cultures were grown to mid-exponential phase (OD600 = 0.6) and total RNA was extracted using Direct-zol RNA Miniprep kit (Zymo Research) according to manufacturer’s instructions. Cells were lysed using Bertin Precellys 24 homogenizer set at 6000 rpm for 40 seconds. Samples were treated with TURBO DNase (Thermo Fisher) followed by clean up with the RNA clean and concentrator kit (Zymo Research) according to manufacturer’s instructions. The absence of DNA contamination was checked by PCR and RNA integrity and purity was checked with a Bioanalyser RNA kit (Agilent). Five sequencing libraries from independent RNA extractions were made for each of the VREfm strains using the Stranded Total RNA with Ribo-Zero Plus (Illumina) kit and sequenced on a single lane of an Illumina NovaSeq 6000 platform. Raw paired-end reads were quality trimmed using TrimGalore (https://www.bioinformatics.babraham.ac.uk/projects/trim_galore/) (v0.6.2). Bases with a quality score <20 and reads shorter than 50bp after trimming were discarded. rRNA was removed by the BBDuk script in BBtools (https://sourceforge.net/projects/bbmap/) (v39.01). The resulting reads were aligned to the Aus0233 reference genome by Bowtie2^74^ (v2.5.1) using the --no-mixed flag and read counts were generated using htseq-count^75^ (v. 0.12.4) using the options -r pos -t CDS -m union --nonunique none. Differentially expressed genes were detected using Degust (v4.1.1). Genes with log_2_(fold change) >1.5 and adjusted p-value < 0.05 were considered differentially expressed.

### Proteomic analysis

Pelleted snap frozen bacterial cells (OD600 = 0.6) were solubilized in 4% SDS, 100mM Tris pH 8.5 by heating them for 10 min at 95 °C. The protein concentrations were assessed by a bicinchoninic acid protein assay (Thermo Fisher Scientific) and 100 µg of each biological replicate prepared for digestion using Micro S-traps (Protifi, USA) according to the manufacturer’s instructions. Briefly, samples were reduced with 10mM DTT for 10 mins at 95°C and then alkylated with 40mM IAA in the dark for 1 hour. Samples were acidified to 1.2% phosphoric acid and diluted with seven volumes of S-trap wash buffer (90% methanol, 100 mM tetraethylammonium bromide pH 7.1) before being loaded onto S-traps and washed 3 times with S-trap wash buffer. Samples were then digested with trypsin before being collected by centrifugation after the addition of 100 mM tetraethylammonium bromide, followed by 0.2% formic acid and then 0.2% formic acid / 50% acetonitrile. Samples were dried and further cleaned up using C18 Stage^76,77^ tips to ensure the removal of any particulate matter.

C18 enriched proteome samples were re-suspended in 2% acetonitrile (aq) containing 0.01% trifluoroacetic acid (Buffer A*) and separated using a Vanquish Neo UHPLC (Thermo Fisher Scientific) with a single column chromatography set up composed of a ACQUITY UPLC Peptide BEH C18 Column (300Å, 1.7 µm, 1 mm X 100 mm, Waters Corporation) at a flow rate of 50 μL/min. Proteome samples were loaded directly on to the ACQUITY column with Buffer A (0.1% formic acid, 2% DMSO) coupled directly to an Orbitrap 480™ mass spectrometer (Thermo Fisher Scientific) and the buffer composition altered from 2% Buffer B (0.1% formic acid, 77.9% acetonitrile, 2% DMSO) to 26% B over 70 minutes, then from 26% B to 99% B over 2 minutes and then was held at 99% B for 1.5 minutes. The mass spectrometer was operated in a data-independent mode automatically switching between the acquisition of a single Orbitrap MS scan (370-1050 m/z, maximal injection time of 50 ms, an AGC set to a maximum of 300% and mass resolving power of 120,000 (at m/z 200) and the collection of 16.5 m/z DIA windows between 375 and 1015 m/z (200-2000 m/z, NCE 32%, maximal injection time of 54 ms, an AGC set to 1000% and a mass resolving power of 30,000 (at m/z 200). Identification and label free quantitation (LFQ) analysis were accomplished using Spectronaut (Biognosys, Switzerland) versions 16 (16.0.220606.53000) using directDIA based analysis with minor modifications: Protein LFQ method set to MaxLFQ, single hit proteins excluded, and imputation disabled. Data was searched against the *E. faecium* Aus0004 proteome^46^ (Uniprot accession: UP000007591) with Carbamidomethyl (C) allowed as a fixed modification and Acetyl (Protein N-term) as well as Oxidation (M) allowed as variable modifications. Data outputs from Spectronaut were processed using Perseus (version 1.6.0.7)^78^ with missing values imputed based on the total observed protein intensities with a range of 0.3 σ and a downshift of 1.8 σ. Statistical analysis was undertaken in Perseus using two-tailed unpaired *t*-tests and ANOVAs. Proteins with log_2_(fold change) >1 and adjusted p-value < 0.05 were considered differentially expressed.

### Computational modelling

In predicting the potential effects of substitutions Q473L, G482D, H486Y, and S491F, the full *E. faecium* DNA-dependent RNA polymerase was initially modelled using Advanced Homology Modelling in Maestro (Schrodinger suite). BLAST-pdb was used to identify the *M. tuberculosis* homologue (PDB: 5UHC)^36^ as the template, as it had the best sequence identities across all RNA polymerase subunits. Modelling was carried out based on consensus between sequence alignments from MAFFT-DASH^79^, T-COFFEE^80^, and Clustal-W^81^ (within Maestro), which were manually optimised to minimise sequence gaps. The final RNA polymerase model, bound to rifampicin and the DNA replication fork was next subjected to loop refinement and minimisation, and iteratively assessed for model quality within Maestro.

The modelled structure was used as input within *in silico* biophysical predictors Dynamut2^82^, mmCSM-lig (https://biosig.lab.uq.edu.au/mmcsm_lig/), mmCSM-NA^83^, and mCSM-PPI2^83^, which predicted the effects of mutations Q473L, G482D, H486Y, and S491F on β-subunit stability, and affinities to rifampicin, nucleic acids within the replication fork, and other RNA polymerase subunits, respectively. During interpretation, all values were collectively considered to assess potential protein-level implications to wild type function. In doing so, the affinity values for mutations located beyond 12 Å of the binding partner were presumed negligible.

### Estimation of Zeta potential

The Zeta potential was measured on cells grown to exponential phase (OD600 = 0.6) and washed in PBS. The Zeta potential measurements were performed in PBS to minimise influence of pH. Each experiment was carried out under identical experiment conditions (n=5), 25°C with two minutes of equilibration. The Zeta potential was measured with a Zetasizer (Malvern, UK).

### Determination of cell-associated daptomycin with BoDipy labelling

BoDipy fluorescent dye (4,4-Difluoro-1,3,5,7,8-Pentamethyl-4-Bora-3a,4a-Diaza-S-Indacene) (Invitrogen) was used to label daptomycin with minor modifications^84^. Briefly, 50 μL daptomycin (50 mg mL^-1^) was mixed with 100 μL BoDipy (10 mg mL^-1^) and was made up to a final volume of 1 mL in 200 mM sodium bicarbonate (pH 8.5). The reaction was incubated for 1 hour at 37°C and unbound BoDipy was removed by dialysis at 4°C using a Slide-A-Lyzer cassette (Thermo Scientific), with a 2.0 kDa cut-off, as per manufacturer’s instructions. The antibiotic activity of BoDipy-daptomycin was confirmed by BMD (described above). To measure cell-associated DAP, cultures were grown to exponential phase (OD600 = 0.6) 50 mg L^-1^ CaCl_2_. Each culture was incubated with BoDipy-daptomycin in darkness (10 minutes) and washed to remove unbound BoDipy-daptomycin. The amount of bound BoDipy-daptomycin was measured with excitation at 490 nm and emission at 528 nm using an Ensight microplate reader (PerkinElmer). Biological replicates (n=5) were completed on separate days.

### Data visualisation and statistics

All figures were generated in R (v4.0.3, https://www.r-project.org/) using tidyverse (v.1.3.1), patchwork (v.1.1.1), and ggnewscale (v.0.4.5). Statistical analyses were performed using R (v4.0.3, https://www.r-project.org/) and GraphPad Prism (v9.3.1) software packages. Specific tests are provided together with each corresponding result in the text.

## Data Availability

The data presented in the study are deposited under Bioprojects PRJNA565795, PRJNA433676, PRJNA856406, and PRJNA856406.

## Ethics approval

Data were obtained from medical records with approval from the local Human Research Ethics Committee (HREC/92972/Austin-2023).

## Competing Interests

The authors declare no competing interests.

## Acknowledgements

This work was supported by the National Health and Medical Research Council (NHMRC) of Australia (GNT1185213 and GNT1160745). BPH is supported by an NHMRC Investigator Grant (GNT1196103). JCK is supported by an NHMRC Early Career Fellowship (GNT1142613). AMT and NLS are supported by an Australian Government Research Training Program scholarship. DJI is supported by an NHMRC Investigator Grant (GNT1195210). SD is supported by the Australian Research Council (DE190100805). The Controlling Superbugs study was supported by the Melbourne Genomics Health Alliance (funded by the State Government of Victoria, Department of Health and Human Services, and the 10 member organizations). NES is supported by an Australian Research Council Future Fellowship (FT200100270) and an ARC Discovery Project Grant (DP210100362). We thank the Melbourne Mass Spectrometry and Proteomics Facility of The Bio21 Molecular Science and Biotechnology Institute for access to MS instrumentation and staff within the Microbiological Diagnostic Unit Public Health Laboratory for technical assistance with antimicrobial susceptibility testing and whole genome sequencing.

## Author Contributions

GPC, BPH, and CLG conceived and planned the experiments. AMT, LL, IRM, DJI, SD, SP, NI, LKS, GR, SN, NES, BAE, DA, and GPC performed the planned experiments. JCK, NEH, BK, DW, AG and EH provided access to necessary patient metadata and JCK and SV preformed statistical analyses in the patient cohort studies. AH, JYHL, NLS, TPS, TS, BPH, EH and JCK provided critical clinical or bioinformatic insights for the study. AMT, CLG, and GPC co-wrote the manuscript with critical feedback and input from all authors.

## Corresponding Authors

Correspondence to Glen Carter or Benjamin Howden.

## Data Availability

The WGS and RNA sequencing data presented in the study are deposited under Bioprojects PRJNA565795, PRJNA433676, PRJNA856406, and PRJNA856406. The mass spectrometry proteomics data has been deposited in the Proteome Xchange Consortium via the PRIDE partner repository^85^ with the data set identifier: PXD039832 (Username: Password: fwjEy0ah) and PXD039831 (Username: Password: 9SrIrmu8).

## Notes

### Competing Interest Statement

The authors have declared no competing interest.

### Summary of Updates

Updated manuscript after reviewer comments.

## References

1. Murray, C. J. et al. Global burden of bacterial antimicrobial resistance in 2019: a systematic analysis. The Lancet 399, 629–655 (2022).

2. Bylicka-Szczepanowska, E. & Korzeniewski, K. Asymptomatic Malaria Infections in the Time of COVID-19 Pandemic: Experience from the Central African Republic. Int. J. Environ. Res. Public. Health 19, 3544 (2022).

3. Global HIV & AIDS statistics — Fact sheet. https://www.unaids.org/en/resources/fact-sheet.

4. GBD 2019 Collaborators. Global mortality from dementia: Application of a new method and results from the Global Burden of Disease Study 2019. Alzheimers Dement. N. Y. N 7, e12200 (2021).

5. Arias, C. A. & Murray, B. E. The rise of the *Enterococcus*: beyond vancomycin resistance. Nat. Rev. Microbiol. 10, 266–278 (2012).

6. Top, J., Willems, R. & Bonten, M. Emergence of CC17 *Enterococcus faecium*: from commensal to hospital-adapted pathogen. FEMS Immunol. Med. Microbiol. 52, 297–308 (2008).

7. Arthur, M. & Courvalin, P. Genetics and mechanisms of glycopeptide resistance in enterococci. Antimicrob. Agents Chemother. 37, 1563–1571 (1993).

8. De Oliveira, D. M. P. et al. Antimicrobial Resistance in ESKAPE Pathogens. Clin. Microbiol. Rev. 33, e00181–19 (2020).

9. Tacconelli, E. et al. Discovery, research, and development of new antibiotics: the WHO priority list of antibiotic-resistant bacteria and tuberculosis. Lancet Infect. Dis. 18, 318– 327 (2018).

10. Montero, C. I., Stock, F. & Murray, P. R. Mechanisms of Resistance to Daptomycin in *Enterococcus faecium*. Antimicrob. Agents Chemother. (2008) doi:10.1128/AAC.00774-07.

11. Diaz, L. et al. Whole-Genome Analyses of *Enterococcus faecium* Isolates with Diverse Daptomycin MICs. Antimicrob. Agents Chemother. (2014).

12. Lellek, H. et al. Emergence of daptomycin non-susceptibility in colonizing vancomycin-resistant *Enterococcus faecium* isolates during daptomycin therapy. Int. J. Med. Microbiol. 305, 902–909 (2015).

13. Kelesidis, T., Tewhey, R. & Humphries, R. M. Evolution of high-level daptomycin resistance in *Enterococcus faecium* during daptomycin therapy is associated with limited mutations in the bacterial genome. J. Antimicrob. Chemother. 68, 1926–1928 (2013).

14. Werth, B. J. et al. Defining Daptomycin Resistance Prevention Exposures in Vancomycin-Resistant *Enterococcus faecium* and *E. faecalis*. Antimicrob. Agents Chemother. 58, 5253–5261 (2014).

15. Turner, A. M., Lee, J. Y. H., Gorrie, C. L., Howden, B. P. & Carter, G. P. Genomic Insights Into Last-Line Antimicrobial Resistance in Multidrug-Resistant *Staphylococcus* and Vancomycin-Resistant *Enterococcus*. Front. Microbiol. 12, (2021).

16. Li, L. et al. Daptomycin Resistance Occurs Predominantly in vanA-Type Vancomycin-Resistant *Enterococcus faecium* in Australasia and Is Associated With Heterogeneous and Novel Mutations. Front. Microbiol. 12, 749935 (2021).

17. Bass, N. M. et al. Rifaximin Treatment in Hepatic Encephalopathy. N. Engl. J. Med. 362, 1071–1081 (2010).

18. Shayto, R. H., Abou Mrad, R. & Sharara, A. I. Use of rifaximin in gastrointestinal and liver diseases. World J. Gastroenterol. 22, 6638–6651 (2016).

19. Sherry, N. L. et al. Pilot study of a combined genomic and epidemiologic surveillance program for hospital-acquired multidrug-resistant pathogens across multiple hospital networks in Australia. Infect. Control Hosp. Epidemiol. 42, 573–581 (2021).

20. Gorrie, C. L. et al. Key parameters for genomics-based real-time detection and tracking of multidrug-resistant bacteria: a systematic analysis. Lancet Microbe 2, e575–e583 (2021).

21. Miller, W. R., Bayer, A. S. & Arias, C. A. Mechanism of Action and Resistance to Daptomycin in *Staphylococcus aureus* and Enterococci. Cold Spring Harb. Perspect. Med. 6, a026997 (2016).

22. Merker, M. et al. Transcontinental spread and evolution of *Mycobacterium tuberculosis* W148 European/Russian clade toward extensively drug resistant tuberculosis. Nat. Commun. 13, 5105 (2022).

23. Brandis, G., Wrande, M., Liljas, L. & Hughes, D. Fitness-compensatory mutations in rifampicin-resistant RNA polymerase. Mol. Microbiol. 85, 142–151 (2012).

24. Goldstein, B. P. Resistance to rifampicin: a review. J. Antibiot. (Tokyo) 67, 625–630 (2014).

25. Rios, R. et al. Genomic Epidemiology of Vancomycin-Resistant *Enterococcus faecium* (VREfm) in Latin America: Revisiting The Global VRE Population Structure. Sci. Rep. 10, 5636 (2020).

26. Duchêne, S. et al. Genome-scale rates of evolutionary change in bacteria. Microb. Genomics 2, e000094 (2016).

27. Lebreton, F. et al. Emergence of Epidemic Multidrug-Resistant *Enterococcus faecium* from Animal and Commensal Strains. mBio 4, e00534–13.

28. Raven, K. E. et al. A decade of genomic history for healthcare-associated *Enterococcus faecium* in the United Kingdom and Ireland. Genome Res. 26, 1388–1396 (2016).

29. Sensi, P. History of the Development of Rifampin. Rev. Infect. Dis. 5, S402–S406 (1983).

30. Goel, A., Rahim, U., Nguyen, L. H., Stave, C. & Nguyen, M. H. Systematic review with meta-analysis: rifaximin for the prophylaxis of spontaneous bacterial peritonitis. Aliment. Pharmacol. Ther. 46, 1029–1036 (2017).

31. Lee, R. A. et al. Daptomycin-Resistant *Enterococcus* Bacteremia Is Associated With Prior Daptomycin Use and Increased Mortality After Liver Transplantation. Open Forum Infect. Dis. 9, ofab659 (2022).

32. Lee, R. S. et al. The changing landscape of vancomycin-resistant *Enterococcus faecium* in Australia: a population-level genomic study. J. Antimicrob. Chemother. 73, 3268–3278 (2018).

33. Weber, D. et al. Rifaximin preserves intestinal microbiota balance in patients undergoing allogeneic stem cell transplantation. Bone Marrow Transplant. 51, 1087–1092 (2016).

34. Taur, Y. et al. Intestinal Domination and the Risk of Bacteremia in Patients Undergoing Allogeneic Hematopoietic Stem Cell Transplantation. Clin. Infect. Dis. 55, 905–914 (2012).

35. Morley, V. J. et al. An adjunctive therapy administered with an antibiotic prevents enrichment of antibiotic-resistant clones of a colonizing opportunistic pathogen. eLife 9, e58147 (2020).

36. Lin, W. et al. Structural Basis of *Mycobacterium tuberculosis* Transcription and Transcription Inhibition. Mol. Cell 66, 169–179.e8 (2017).

37. Portelli, S., Phelan, J. E., Ascher, D. B., Clark, T. G. & Furnham, N. Understanding molecular consequences of putative drug resistant mutations in *Mycobacterium tuberculosis*. Sci. Rep. 8, 15356 (2018).

38. Bender, J. K. et al. Update on prevalence and mechanisms of resistance to linezolid, tigecycline and daptomycin in enterococci in Europe: Towards a common nomenclature. Drug Resist. Updat. 40, 25–39 (2018).

39. Tran, T. T. et al. Whole-Genome Analysis of a Daptomycin-Susceptible *Enterococcus faecium* Strain and Its Daptomycin-Resistant Variant Arising during Therapy. Antimicrob. Agents Chemother. 57, 261–268 (2013).

40. Guérillot, R. et al. Convergent Evolution Driven by Rifampin Exacerbates the Global Burden of Drug-Resistant *Staphylococcus aureus*. mSphere 3, e00550-17.

41. Bæk, K. T. et al. Stepwise Decrease in Daptomycin Susceptibility in Clinical *Staphylococcus aureus* Isolates Associated with an Initial Mutation in *rpoB* and a Compensatory Inactivation of the *clpX* Gene. Antimicrob. Agents Chemother. (2015).

42. Cui, L. et al. An RpoB Mutation Confers Dual Heteroresistance to Daptomycin and Vancomycin in *Staphylococcus aureus*. Antimicrob. Agents Chemother. (2010) doi:10.1128/AAC.00437-10.

43. Snyder, E. E. et al. PATRIC: The VBI PathoSystems Resource Integration Center. Nucleic Acids Res. 35, D401–D406 (2007).

44. Wood, D. E., Lu, J. & Langmead, B. Improved metagenomic analysis with Kraken 2. Genome Biol. 20, 257 (2019).

45. Humphries, R. M. The New, New Daptomycin Breakpoint for *Enterococcus* spp. J. Clin. Microbiol. 57, e00600–19.

46. Lam, M. M. et al. Comparative analysis of the complete genome of an epidemic hospital sequence type 203 clone of vancomycin-resistant *Enterococcus faecium*. BMC Genomics 14, 595 (2013).

47. Bankevich, A. et al. SPAdes: A New Genome Assembly Algorithm and Its Applications to Single-Cell Sequencing. J. Comput. Biol. 19, 455–477 (2012).

48. Homan, W. L. et al. Multilocus sequence typing scheme for *Enterococcus faecium*. J. Clin. Microbiol. 40, 1963–1971 (2002).

49. Minh, B. Q. et al. IQ-TREE 2: New Models and Efficient Methods for Phylogenetic Inference in the Genomic Era. Mol. Biol. Evol. 37, 1530–1534 (2020).

50. Schliep, K. P. phangorn: phylogenetic analysis in R. Bioinformatics 27, 592–593 (2011).

51. Paradis, E. & Schliep, K. ape 5.0: an environment for modern phylogenetics and evolutionary analyses in R. Bioinformatics 35, 526–528 (2019).

52. Yu, G., Smith, D. K., Zhu, H., Guan, Y. & Lam, T. T.-Y. ggtree: an r package for visualization and annotation of phylogenetic trees with their covariates and other associated data. Methods Ecol. Evol. 8, 28–36 (2017).

53. Sherry, N. L. et al. An ISO-certified genomics workflow for identification and surveillance of antimicrobial resistance. Nat. Commun. 14, 60 (2023).

54. Crispell, J., Balaz, D. & Gordon, S. V. HomoplasyFinder: a simple tool to identify homoplasies on a phylogeny. Microb. Genomics 5, e000245 (2019).

55. Lees, J. A., Galardini, M., Bentley, S. D., Weiser, J. N. & Corander, J. pyseer: a comprehensive tool for microbial pangenome-wide association studies. Bioinformatics 34, 4310–4312 (2018).

56. de Been, M. et al. Core Genome Multilocus Sequence Typing Scheme for High-Resolution Typing of *Enterococcus faecium*. J. Clin. Microbiol. 53, 3788–3797 (2015).

57. Silva, M. et al. chewBBACA: A complete suite for gene-by-gene schema creation and strain identification. Microb. Genomics 4, e000166 (2018).

58. Croucher, N. J. et al. Rapid phylogenetic analysis of large samples of recombinant bacterial whole genome sequences using Gubbins. Nucleic Acids Res. 43, e15 (2015).

59. Nguyen, L.-T., Schmidt, H. A., von Haeseler, A. & Minh, B. Q. IQ-TREE: A Fast and Effective Stochastic Algorithm for Estimating Maximum-Likelihood Phylogenies. Mol. Biol. Evol. 32, 268–274 (2015).

60. Rambaut, A., Lam, T. T., Max Carvalho, L. & Pybus, O. G. Exploring the temporal structure of heterochronous sequences using TempEst (formerly Path-O-Gen). Virus Evol. 2, vew007 (2016).

61. Suchard, M. A. et al. Bayesian phylogenetic and phylodynamic data integration using BEAST 1.10. Virus Evol. 4, vey016 (2018).

62. Minin, V. N. & Suchard, M. A. Counting labeled transitions in continuous-time Markov models of evolution. J. Math. Biol. 56, 391–412 (2008).

63. Minin, V. N. & Suchard, M. A. Fast, accurate and simulation-free stochastic mapping. Philos. Trans. R. Soc. B Biol. Sci. 363, 3985–3995 (2008).

64. Monk, I. R., Tree, J. J., Howden, B. P., Stinear, T. P. & Foster, T. J. Complete Bypass of Restriction Systems for Major *Staphylococcus aureus* Lineages. mBio 6, e00308–15.

65. Zhang, Y., Werling, U. & Edelmann, W. Seamless Ligation Cloning Extract (SLiCE) Cloning Method. in DNA Cloning and Assembly Methods (eds. Valla, S. & Lale, R.) 235–244 (Humana Press, Totowa, NJ, 2014). doi:10.1007/978-1-62703-764-8_16.

66. Pidot, S. J. et al. Increasing tolerance of hospital *Enterococcus faecium* to handwash alcohols. Sci. Transl. Med. 10, eaar6115 (2018).

67. Nair, A. B. & Jacob, S. A simple practice guide for dose conversion between animals and human. J. Basic Clin. Pharm. 7, 27–31 (2016).

68. Heine, H. S., Bassett, J., Miller, L., Purcell, B. K. & Byrne, W. R. Efficacy of Daptomycin against *Bacillus anthracis* in a Murine Model of Anthrax Spore Inhalation. Antimicrob. Agents Chemother. 54, 4471–4473 (2010).

69. Higgs, C. et al. Optimising genomic approaches for identifying vancomycin-resistant *Enterococcus faecium* transmission in healthcare settings. Nat. Commun. 13, 509 (2022).

70. Maechler, F. et al. Split k-mer analysis compared to cgMLST and SNP-based core genome analysis for detecting transmission of vancomycin-resistant enterococci: results from routine outbreak analyses across different hospitals and hospitals networks in Berlin, Germany. Microb. Genomics 9, 000937 (2023).

71. Liese, J. et al. Expansion of Vancomycin-Resistant *Enterococcus faecium* in an Academic Tertiary Hospital in Southwest Germany: a Large-Scale Whole-Genome-Based Outbreak Investigation. Antimicrob. Agents Chemother. 63, e01978–18 (2019).

72. Kralj, T. et al. Multi-Omic Analysis to Characterize Metabolic Adaptation of the *E. coli* Lipidome in Response to Environmental Stress. Metabolites 12, (2022).

73. Liebisch, G. et al. Update on LIPID MAPS classification, nomenclature, and shorthand notation for MS-derived lipid structures. J. Lipid Res. 61, 1539–1555 (2020).

74. Langmead, B. & Salzberg, S. L. Fast gapped-read alignment with Bowtie 2. Nat. Methods 9, 357–359 (2012).

75. Anders, S., Pyl, P. T. & Huber, W. HTSeq—a Python framework to work with high-throughput sequencing data. Bioinformatics 31, 166–169 (2015).

76. Rappsilber, J., Ishihama, Y. & Mann, M. Stop and Go Extraction Tips for Matrix-Assisted Laser Desorption/Ionization, Nanoelectrospray, and LC/MS Sample Pretreatment in Proteomics. Anal. Chem. 75, 663–670 (2003).

77. Rappsilber, J., Mann, M. & Ishihama, Y. Protocol for micro-purification, enrichment, pre-fractionation and storage of peptides for proteomics using StageTips. Nat. Protoc. 2, 1896–1906 (2007).

78. Cox, J. & Mann, M. MaxQuant enables high peptide identification rates, individualized p.p.b.-range mass accuracies and proteome-wide protein quantification. Nat. Biotechnol. 26, 1367–1372 (2008).

79. Rozewicki, J., Li, S., Amada, K. M., Standley, D. M. & Katoh, K. MAFFT-DASH: integrated protein sequence and structural alignment. Nucleic Acids Res. 47, W5–W10 (2019).

80. Magis, C. et al. T-Coffee: Tree-based consistency objective function for alignment evaluation. Methods Mol. Biol. Clifton NJ 1079, 117–129 (2014).

81. Higgins, D. G. & Sharp, P. M. CLUSTAL: a package for performing multiple sequence alignment on a microcomputer. Gene 73, 237–244 (1988).

82. Rodrigues, C. H. M., Pires, D. E. V. & Ascher, D. B. DynaMut2: Assessing changes in stability and flexibility upon single and multiple point missense mutations. Protein Sci. Publ. Protein Soc. 30, 60–69 (2021).

83. Nguyen, T. B., Myung, Y., de Sá, A. G. C., Pires, D. E. V. & Ascher, D. B. mmCSM-NA: accurately predicting effects of single and multiple mutations on protein-nucleic acid binding affinity. NAR Genomics Bioinforma. 3, lqab109 (2021).

84. Pader, V., et al. *Staphylococcus aureus* inactivates daptomycin by releasing membrane phospholipids. Nat. Microbiol. 2, 1–8 (2016).

85. Perez-Riverol, Y. et al. The PRIDE database resources in 2022: a hub for mass spectrometry-based proteomics evidences. Nucleic Acids Res. 50, D543–D552 (2022).

